# Prospects of HIV elimination among men who have sex with men: a systematic review of modeling studies

**DOI:** 10.1101/2024.09.22.24313968

**Authors:** Jacob Aiden Roberts, Alexandra Teslya, Mirjam E. Kretzschmar, Janneke H. H. M. van de Wijgert, Ganna Rozhnova

## Abstract

**Background:** Despite advances in HIV treatment and prevention, men who have sex with men (MSM) remain disproportionately affected by HIV worldwide. This systematic review summarizes the results of mathematical modeling studies that evaluated whether interventions might eliminate HIV in MSM populations by geographical setting, type of intervention(s), elimination definition, and model characteristics.

**Methods:** We searched Embase and PubMed for modeling studies published between July 1, 2016 and January 7, 2025. Studies were included if they used a dynamic model to assess the impact of interventions on HIV transmission among MSM. Data were extracted on article information, study population, interventions, elimination definitions, model type, model structure, and calibration. The studies were critically appraised by evaluating the comprehensiveness of their models in addressing elimination.

**Findings:** Of the 3,250 identified records, 89 studies were included. MSM populations in only five of the eight Joint United Nations Programme on HIV/AIDS (UNAIDS) regions were modeled, with over half of the models considering MSM in the USA. Complex agent-based models (ABMs) were as common as simpler compartmental models overall, with ABMs more frequently used in Western and Central Europe and North America (WCENA), while compartmental models predominated elsewhere. Thirty-nine of the 89 studies defined elimination as reductions or thresholds in HIV incidence or prevalence, a reproduction number below one, or the elimination of racial disparities. Elimination was achieved in 36 out of 50 modeled scenarios, but the authors of only six of these 36 scenarios thought the interventions required to achieve elimination were feasible. The six feasible elimination scenarios were reported in compartmental models for few countries in Western Europe and Asia. Models in which elimination was achieved most commonly used a combination of interventions that included pre-exposure prophylaxis (PrEP) and/or test-and-treat, except in Africa, where PrEP was not included.

**Interpretation:** Modeling efforts to understand HIV elimination prospects among MSM outside WCENA should be intensified. To enhance study comparability and for models to contribute effectively to public health policy, the use of an elimination definition based on an incidence threshold would be the most valuable. Furthermore, by identifying gaps in current studies, we recommend novel research directions for modeling to inform a coordinated global response for HIV elimination among MSM.

## Introduction

The HIV epidemic remains a major public health problem, particularly among key populations [1]. The Joint United Nations Programme on HIV/AIDS (UNAIDS) considers men who have sex with men (MSM) as one of the main key populations vulnerable to HIV acquisition and transmission. Despite considerable progress in HIV prevention and treatment overall, MSM continue to have a disproportionately high HIV incidence and prevalence worldwide [1]. In 2022, global HIV prevalence among MSM was eleven times higher than among adults in the general population [2]. Alarmingly, the annual number of new HIV infections among MSM increased by 11% globally and by 19% outside sub-Saharan Africa from 2010 to 2022 [3]. Contrasting trends in reaching elimination among MSM are currently observed in different UNAIDS regions. Some countries in Western and Central Europe and North America are characterized by a rapidly slowing epidemic (e.g., [4–6]), while emerging and ongoing HIV epidemics are reported in the Middle East and North Africa [7, 8], the Caribbean [9], and no evidence of slowing epidemics is found in Africa [10].

To address a disproportionate burden of HIV among MSM, interventions such as classical partner reduction and condom use approaches, pre-exposure prophylaxis (PrEP) [11], and test-and-treat [12] are used either separately or in combination to reach this key population [13]. Predicting the impact of interventions on HIV dynamics at the population level empirically is challenging. Mathematical modeling can guide the design of interventions and plays an increasingly important role in supporting evidence-based policymaking in public health [14]. Models describe transmission dynamics using equations and/or computer simulations. Modeling studies are often the only way to investigate large-scale complex HIV dynamics, particularly in cases where experiments are not ethical or logistically impossible. A well-designed model can assist policymakers in making decisions on HIV control and elimination. Despite a large body of modeling studies that investigate the impact of interventions on HIV transmission dynamics [15], including the assessment of HIV elimination strategies [4, 16], the literature concerning the prospects of HIV elimination among MSM worldwide is inconsistent. The success of HIV elimination in a specific context depends on a combination of factors, such as the target population, the state of the HIV epidemic, HIV care and prevention practices, and details of sexual behavior that shape HIV transmission among MSM. Evaluation of HIV elimination prospects is complicated by the fact that different authors use different definitions of elimination, posing a barrier to a unified response. HIV elimination is usually considered accomplished upon reaching a certain quantifiable threshold, often based on guidelines issued by (inter)national public health authorities. Definitions of elimination include achieving zero new HIV infections, reducing HIV incidence to a low level, or reaching a point where the HIV epidemic is no longer a public health threat [17]. A 90% reduction in HIV incidence by 2030 is an example of the latter definition used in ending the HIV epidemic goals in the USA [18]. The elimination definition of fewer than one HIV infection per 1,000 persons per year was adopted from the seminal modeling study by Granich et al. [12], which received attention from the public health community by demonstrating the possibility of HIV elimination. This systematic review aims to improve our understanding of the prospects of HIV elimination among MSM globally. We summarized mathematical modeling studies by (i) geographical setting where elimination may or may not be achieved, (ii) elimination definitions used, and (iii) interventions required to achieve elimination. We discuss the knowledge gaps in these areas and identify further modeling research needed to better inform policy about effective intervention strategies for ultimately achieving the ambitious goal of HIV elimination among MSM.

## Methods

This systematic review adhered to PRISMA guidelines [19]. The full PRISMA checklist is available in Appendix 1. No protocol was registered for this review.

### Search strategy and selection criteria

Embase and PubMed were searched for studies published between July 1, 2016 and January 7, 2025, when the search was conducted. The starting date was chosen to coincide with the publication of the World Health Organization (WHO) consolidated guidelines on the use of antiretroviral drugs for treating and preventing HIV infection [20]. The search string was (“HIV” OR “human immunodeficiency virus”) AND (“homosexual*” OR “transgender*” OR “gay” OR “MSM” OR “men who have sex with men” OR “men having sex with men” OR “bisexual*”) AND (“model*” OR “framework” OR “simulat*”) AND (“treat*” OR “prevent*”) AND (“mathematic*” OR “transm*” OR “comput*”).

Studies were included if they (i) involved a dynamic model for HIV transmission, where the force of infection depends on the state of the population at a given time, and (ii) assessed the impact of interventions on HIV transmission among MSM. Studies for HIV transmission in a broader population involving MSM were included if they reported a direct or indirect impact of interventions on HIV outcomes among MSM specifically. Studies that involved a dynamic co-transmission model of HIV and another sexually transmitted infection (STI) were included if the primary outcome was HIV. As we were interested in the epidemiological impact of interventions assessed using dynamic transmission models, statistical, back-calculation, decision-analytic, and health economic models primarily focused on cost-effectiveness evaluations were excluded. Studies focused on methodology rather than the impact of interventions on HIV transmission were excluded. Conference abstracts, reviews, preprints, articles not published in English, or without full text were also excluded.

### Data extraction and analysis

Two authors independently screened the titles and abstracts for inclusion and identified eligible studies using Rayyan software. Three authors independently conducted full-text screening and extracted data using a predefined data extraction form. Study inclusion was by consensus. Rare disagreements were resolved through detailed discussions of the studies in question. The authors regularly compared extracted data to ensure consistency in the review process. Data were extracted on article information, study population, interventions, elimination definition, model type, model structure, and calibration (43 data fields in total; Appendix 2). Data fields were summarized descriptively unless quantitative data were available.

Studies were categorized geographically by country and by UNAIDS region (Asia and the Pacific, Caribbean, Eastern and Southern Africa, Eastern Europe and Central Asia, Latin America, Middle East and North Africa, Western and Central Africa, Western and Central Europe and North America) [2]. Additionally, studies were categorized by characteristics of the modeled population, such as demographics (age and ethnicity), subgroups (MSM only or MSM and other subgroups such as the heterosexual population, female sex workers and their clients, injecting drug users, and transgender women), and geographical scale (national, regional, or urban). National scale models consider populations in the entire country. Regional scale models consider a large administrative division within a country (e.g., a state in the USA or a province in China). Urban scale models consider a city or a group of cities and their immediate metropolitan areas. Populations were regarded as stratified by these characteristics if model analyses used different model parameters to describe distinct subgroups.

Models were categorized into deterministic compartmental (DCM), stochastic compartmental (SCM), and agent-based models (ABM). Compartmental models stratify the population into compartments based on certain characteristics, such as disease stage, and track the population in each compartment over time. In contrast, agent-based models include individual heterogeneities and track the status of each individual over time. Unlike deterministic models, stochastic models account for random events. Agent-based models are inherently stochastic.

We reported primary interventions, defined as interventions for which model parameters describing different aspects of intervention engagement (e.g., uptake, retention, coverage, and adherence) and/or intervention efficacy were varied. For example, in a study investigating the increase in PrEP uptake combined with regular HIV testing, PrEP is considered the primary intervention because its uptake was varied. The definition of HIV elimination was a measurable target used in the model analyses to determine when interventions could stop HIV transmission. Elimination definitions and interventions required to achieve elimination were extracted and narratively reported. Descriptive statistics, i.e., frequencies and percentages, were used to summarize the study locations, characteristics of modeled populations, elimination definitions used, and interventions evaluated. Summary statistics were presented in tables and bar charts. The locations of the studied populations and HIV prevalence were visualized using world maps. Subgroup analyses of the summary statistics were performed to examine differences in elimination prospects, stratified by UNAIDS region, model type, elimination definition, and interventions. All results were pooled quantitatively whenever feasible.

### Critical appraisal

Studies that included an elimination definition were critically appraised by evaluating the comprehensiveness of the models in addressing elimination. Given that different studies pursued different goals, ranging from conceptual analytical investigations to operational modeling, the critical appraisal was not used to exclude studies but rather to evaluate the appropriateness of model structures and methodologies for assessing elimination. In the absence of standardized tools for the critical appraisal of modeling studies focused on elimination, we developed our own scoring system. The model comprehensiveness score was calculated based on five criteria, such as whether a model accounted for adherence to interventions, sexual risk compensation, the openness of the modeled MSM population (indicating that new HIV infections could be imported or result from sexual contacts with the external population), whether uncertainty in the model outcomes was investigated and reported, and whether a model was validated. Table S4 outlines the questions we used to assess each criterion and provides clear guidance on scoring. Studies received a score between 0 and 2 for the inclusion of adherence to interventions, reporting of uncertainty in model outcomes, and model validation. A score of 0 or 1 was assigned for the inclusion of sexual risk compensation and the openness of the MSM population. The individual scores were then summed to obtain the final score for each study, ranging from 0 to 8, with a higher score indicating that more criteria were satisfied. Critical appraisal was performed independently by two authors. The scoring of studies was by consensus.

## Results

### Study selection

The initial search resulted in 3,250 records, of which 89 studies (3.94%) met the eligibility criteria and were included for data extraction (Figure 1). After removing duplicates and screening titles and abstracts, 3,048 records (93,78%) were excluded. The majority of records were excluded because they did not involve a dynamic transmission model (*n* = 1, 727, 83.07%). Following the full-text assessment, a further 93 of 182 records (51.10%) were excluded; due to being conference abstracts (*n* = 28, 15.38%), lack of a dynamic transmission model (*n* = 21, 11.54%), or for other reasons (*n* = 49, 26.92%). The list of 89 included studies and their characteristics is given in Table 1.

**Figure 1.**
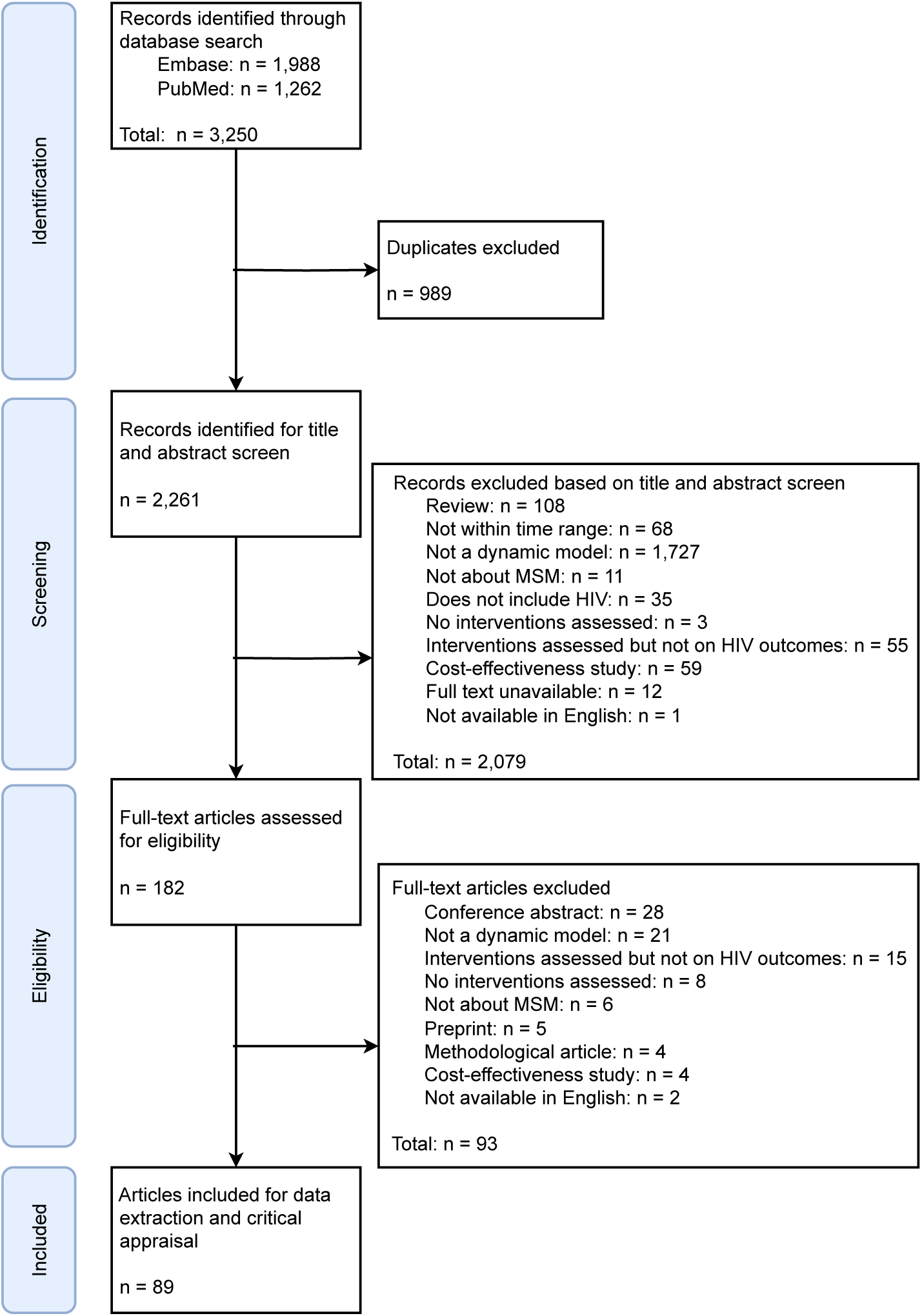
Study selection. MSM = men who have sex with men.

**Table 1.**
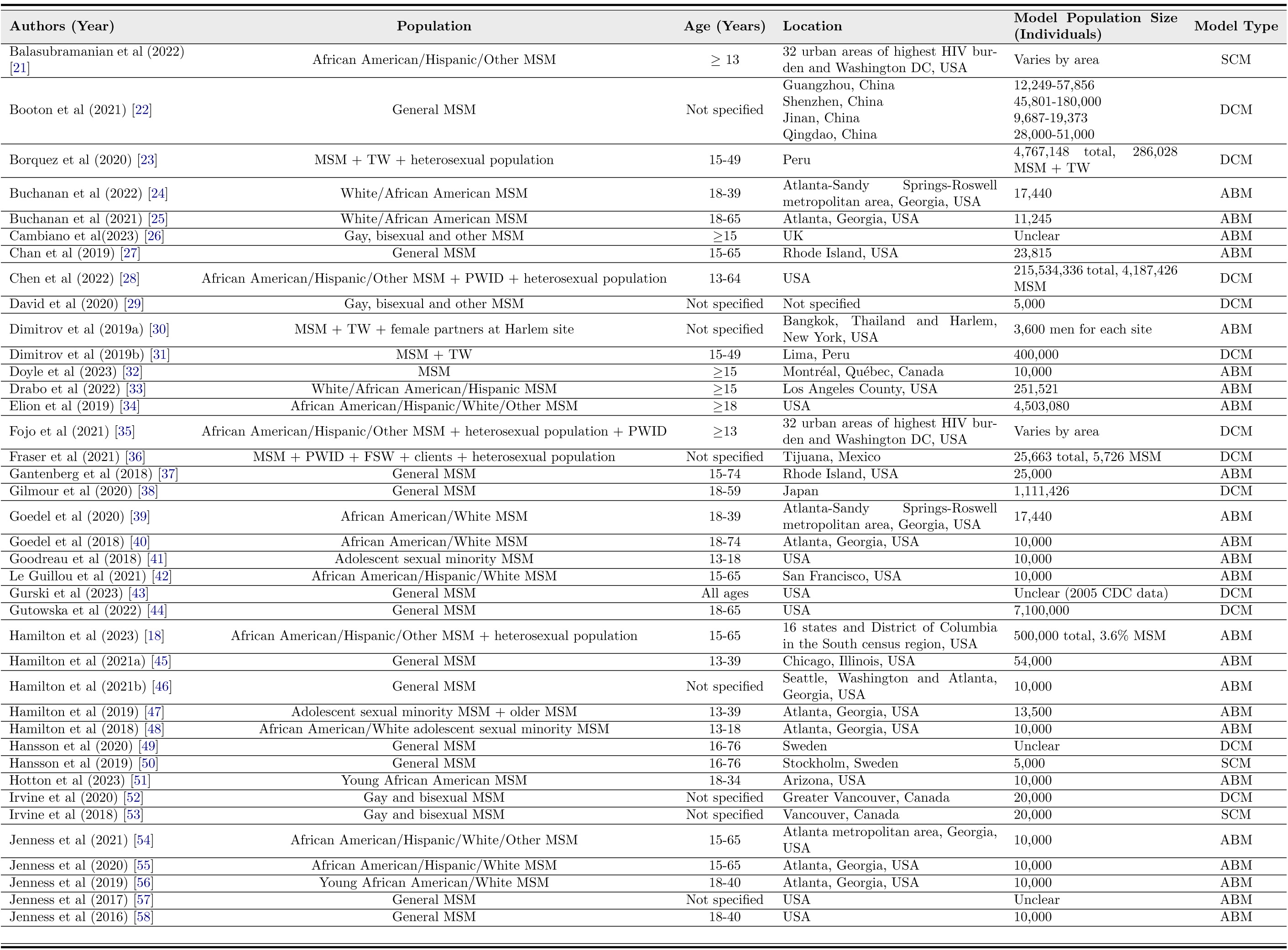
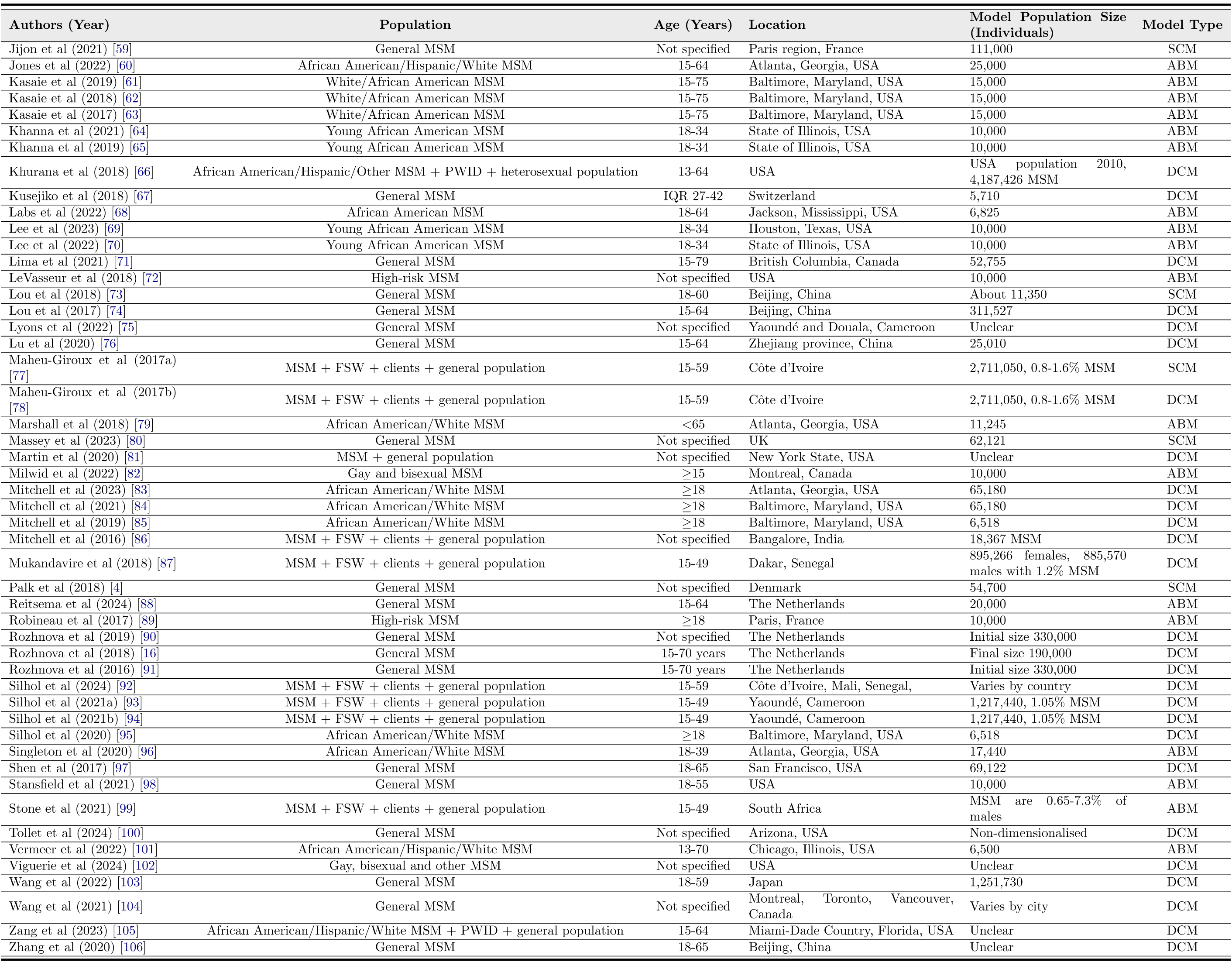
Characteristics of the included studies. MSM = men who have sex with men. TW = transgender women. PWID = people who inject drugs. FSW = female sex workers. SCM = stochastic compartmental model. DCM = deterministic compartmental model. ABM = agent-based model. IQR = interquartile range.

### Study locations and HIV prevalence

Figure 2 shows the worldwide location of the modeled populations, juxtaposed with HIV prevalence among MSM by country. Of the 89 included studies, sixty-nine (77.53%) focused on MSM in Western and Central Europe and North America, nine (10.11%) on Asia and the Pacific, seven (7.87%) on Western and Central Africa, three (3.37%) on Latin America, one (1.12%) on Eastern and Southern Africa, one (1.12%) on two UNAIDS regions [30], and one (1.12%) was not associated with a specified geographical location [29] (Figure 2 **A**). No studies meeting the criteria for our review focused on MSM in the Caribbean, Eastern Europe and Central Asia, or the Middle East and North Africa. More than half of included studies considered MSM populations in the USA (*n* = 51, 57.30%), with the second most frequently considered location being Canada (*n* = 6, 6.74%) (Figure 2 **C**). We observed a striking discordance between the geography of the modeled populations and HIV prevalence among MSM in those populations (Figure 2 **B** and **C**). For example, South Africa, which has the highest HIV prevalence among MSM (29.7%), is represented by a single study (1.12%). Additionally, we found only one study for Senegal (1.12%) and three studies for Cameroon (3.37%), which have the next highest HIV prevalence among MSM (27.6% and 20.6%, respectively). The HIV prevalence in MSM in Western countries, such as Switzerland, France, and the USA, is similar (about 14-15%), however, most studies focus on the USA, with only a few studies considering MSM in other Western countries. Across all UNAIDS regions, 29 countries with HIV prevalence among MSM above 15% [1] were not represented in the studies in this review.

**Figure 2.**
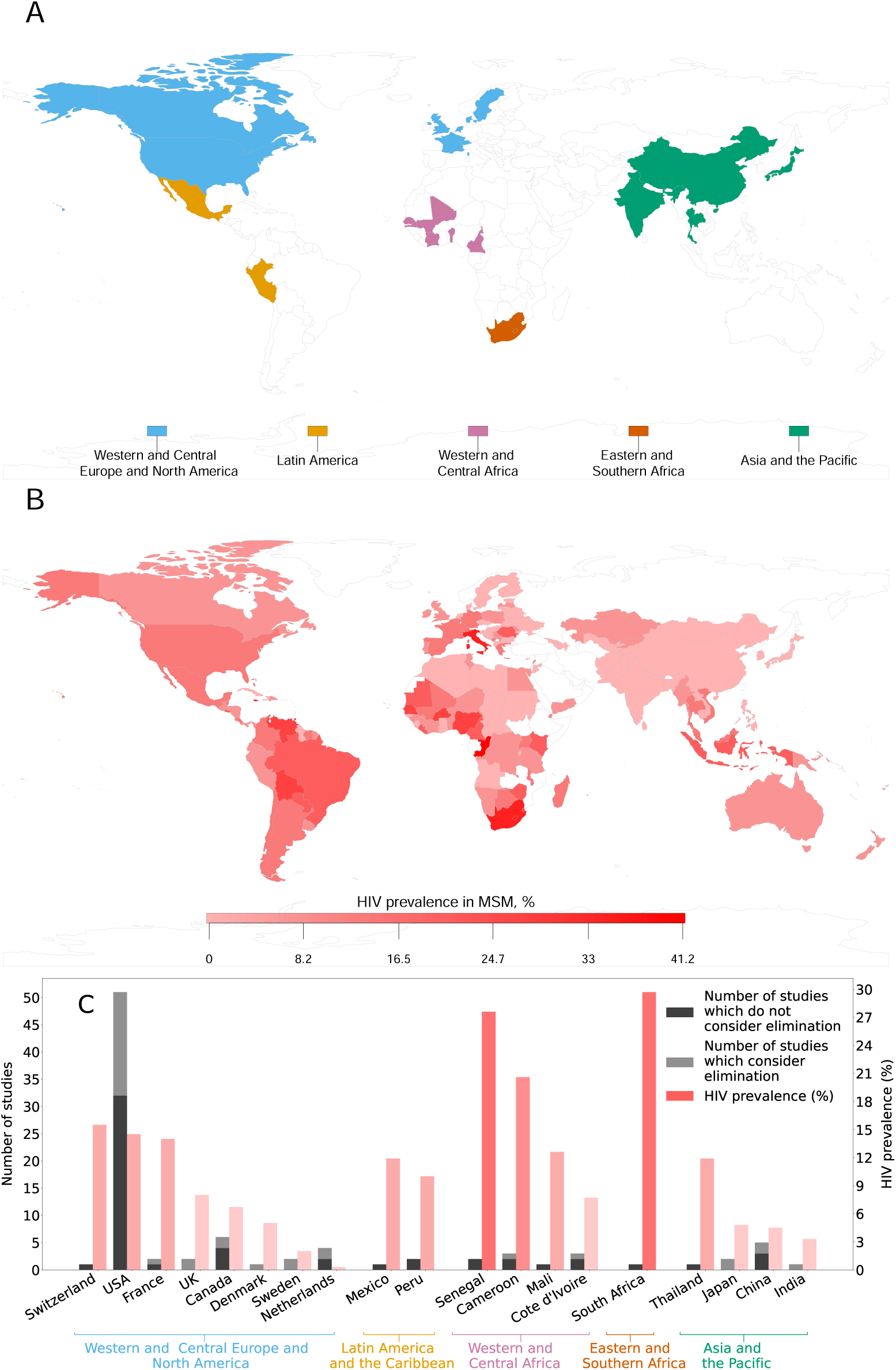
Worldwide location of studies and HIV prevalence among MSM. (**A**) Locations of the modeled populations by UNAIDS region [2]. (**B**) HIV prevalence among MSM by country reported by UNAIDS [1]. In (**A**) and (**B**), countries with no data are shown in white. (**C**) The number of studies that did or did not define elimination and HIV prevalence among MSM by country. The study [30] that considered two countries was counted twice, and the study [92] that considered three countries was counted thrice. The study [29] that did not apply to any specific location was not counted. MSM = men who have sex with men.

### Population characteristics and model types

The distribution of studies by the characteristics of the modeled population and model types across UNAIDS regions is shown in Table 2. All 28 studies (40.58%) in the Western and Central Europe and North America region that stratified MSM by ethnicity concerned MSM populations at different geographical scales in the USA. Commonly considered subgroups were non-Hispanic Black or African American, Hispanic or Latino, non-Hispanic White or other (e.g., [18, 21, 39]). Most studies that included stratification by ethnicity explored the effectiveness of interventions in achieving the dual goals of reducing the overall HIV burden and narrowing racial disparities (e.g., [39, 40, 48]). None of the studies in other regions used stratification by ethnicity. Thirty-two studies (47.83%) in Western and Central Europe and North America, one study (33.33%) in Latin America [31], and six studies (85.71%) in Western and Central Africa [77, 78, 87, 92–94] stratified MSM by age. None of the studies for Asia and the Pacific or Eastern and Southern Africa used age stratification. The age ranges mostly covered sexually active MSM, spanning from 13–18 years to 60–80 years. Two studies, both in USA populations, included adolescent sexual minority men (13 to 18 years old) to investigate the impact of interventions on HIV burden in this group specifically [41, 48]. The majority of studies considered populations consisting solely of MSM (*n* = 69, 76.67%) (e.g., [16, 22, 99, 106]). The remaining studies (*n* = 21, 23.33%) included other subgroups, such as the heterosexual population [18], transgender women [31], injecting drug users, female sex workers and their clients [86]. Most studies for settings in Western and Central Africa (*n* = 6, 85.71%) [75, 77, 78, 87, 92, 93], and Eastern and Southern Africa (*n* = 1, 100%) [99] considered sexual mixing of MSM and the consequent cross-transmission of HIV with other subgroups. Studies for Western countries mostly consider MSM as a separate key population with just a few studies considering MSM mixing with other subgroups (*n* = 7, 10.14%) [16, 18, 28, 30, 66, 81, 105]. Most studies developed models for an urban environment (*n* = 51, 56.67%), while the remaining studies developed national (*n* = 26, 28.89%) and regional (*n* = 12, 13.33%) models, or did not mention any particular geographical scale (*n* = 1, 1.11%) [29]. ABMs were the most frequently used model type overall (*n* = 43, 47.78%) but were rarely used outside Western countries (*n* = 2, 9.52%). DCMs were the second most used model type (*n* = 40, 44.44%), being the main model type in Latin America (*n* = 3, 100.00%), Western and Central Africa (*n* = 6, 85.71%), and Asia and the Pacific (*n* = 7, 77.78%). SCMs were rarely used (*n* = 7, 7.78%).

**Table 2.** Number of studies by characteristics of the modeled population and model types. MSM = men who have sex with men. DCM = deterministic compartmental model. SCM = stochastic compartmental model. ABM = agent-based model. UNAIDS = The Joint United Nations Programme on HIV/AIDS.

| Population characteristics |  | UNAIDS regions* |  |  |  |  | Total |
| --- | --- | --- | --- | --- | --- | --- | --- |
|  |  | Western and Central Europe and North America | Latin America | Western and Central Africa | Eastern and Southern Africa | Asia and The Pacific |  |
|  |  | n (%) | n (%) | n (%) | n (%) | n (%) | n (%) |
| Ethnicity | Yes | 28 (40.58) | 0 (0.00) | 0 (0.00) | 0 (0.00) | 0 (0.00) | **28** (31.11) |
|  | No | 41 (50.42) | 3 (100.00) | 7 (100.00) | 1 (100.00) | 9 (100.00) | **62\*** (68.89) |
| Age | Yes | 33 (47.83) | 1 (33.33) | 6 (85.71) | 0 (0.00) | 0 (0.00) | **40** (44.44) |
|  | No | 36 (52.17) | 2 (66.67) | 1 (14.29) | 1 (100.00) | 9 (100.00) | **50\*** (55.56) |
| MSM only<br>(vs MSM + other subgroups) | Yes | 60 (86.96) | 0 (0.00) | 1 (14.29) | 0 (0.00) | 7 (77.78) | **69\*** (76.67) |
|  | No | 9 (13.04) | 3 (100.00) | 6 (85.71) | 1 (100.00) | 2 (22.22) | **21** (23.33) |
| Geographical scale | National | 19 (27.54) | 1 (33.33) | 3 (42.86) | 1 (100.00) | 2 (22.22) | **26** (28.89) |
|  | Regional | 11 (15.94) | 0 (0.0) | 0 (0.00) | 0 (0.00) | 1 (11.11) | **12** (13.33) |
|  | Urban | 39 (56.52) | 2 (66.67) | 4 (57.14) | 0 (0.00) | 6 (66.67) | **51** (56.67) |
| Model type | DCM | 23 (33.33) | 3 (100.00) | 6 (85.71) | 0 (0.00) | 7 (77.78) | **40\*** (44.44) |
|  | SCM | 5 (7.25) | 0 (0.00) | 1 (14.29) | 0 (0.00) | 1 (11.11) | **7** (7.78) |
|  | ABM | 41 (59.42) | 0 (0.00) | 0 (0.00) | 1 (100.00) | 1 (11.11) | **43** (47.78) |
| **Total** |  | **69** | **3** | **7** | **1** | **9** | **90\*** |

\* The study [29] considered only MSM using a DCM and did not reference any specific location, geographical scale, age, or ethnicity. The study [30] that included both the USA and Thailand was counted for two UNAIDS regions.

### Interventions

Studies investigated classical behavioral interventions (partner reduction and condom use), biomedical interventions (PrEP, post-exposure prophylaxis (PEP), test-and-treat, voluntary medical male circumcision (VMMC), and STI treatment), and structural interventions. Classical behavioral interventions facilitated changes in the behavior of MSM pertinent to HIV transmission, such as a reduction in the number of sexual partners (e.g., [74, 76, 103]), and increased use or effectiveness of condoms (e.g., [36, 43, 77]). The term test-and-treat was used to describe interventions that accelerated HIV testing and/or ART initiation (e.g., [29,85,91]). Structural interventions involved changes in healthcare systems or support for MSM at risk, such as providing housing for homeless MSM [81]. Figure 3 shows the distribution of interventions by type and UNAIDS region. Globally, the most frequently studied interventions were biomedical, namely PrEP and test-and-treat, followed by classical behaviour HIV prevention approaches. In contrast, STI treatment, PEP, VMMC, and structural interventions were rarely included in modeling studies. PrEP was considered more often (*n* = 57, 49.57%) in Western and Central Europe and North America (e.g., [41, 58, 61]) than in other regions (*n* = 1, 33.33% in Latin America [23]; *n* = 4, 21.05% in Asia and the Pacific [30, 38, 86, 103]; *n* = 2, 14.29% in Western and Central Africa [75, 87]). The next most studied intervention,test-and-treat, was considered frequently across all included regions (*n* = 1, 100.00% in Eastern and Southern Africa [99]; *n* = 6, 42.86% in Western and Central Africa [77, 78, 87, 92–94]; *n* = 1, 33.33% in Latin America [36]; *n* = 6, 31.58% in Asia and the Pacific [22, 30, 73, 74, 76, 103]; *n* = 37, 32.17% in Western and Central Europe and North America, e.g., [43, 66, 81]). Classical behavioral interventions were studied frequently outside Western and Central Europe and North America. Combinations of different interventions were more frequently investigated in Western and Central Africa (*n* = 6, 85.71%) [78, 87, 93, 94], Asia and the Pacific (*n* = 7, 77.78%) [22, 30, 38, 73, 74, 76, 103], and Latin America (*n* = 2, 66.67%) [23, 36] than in other regions (Figure S1).

**Figure 3.**
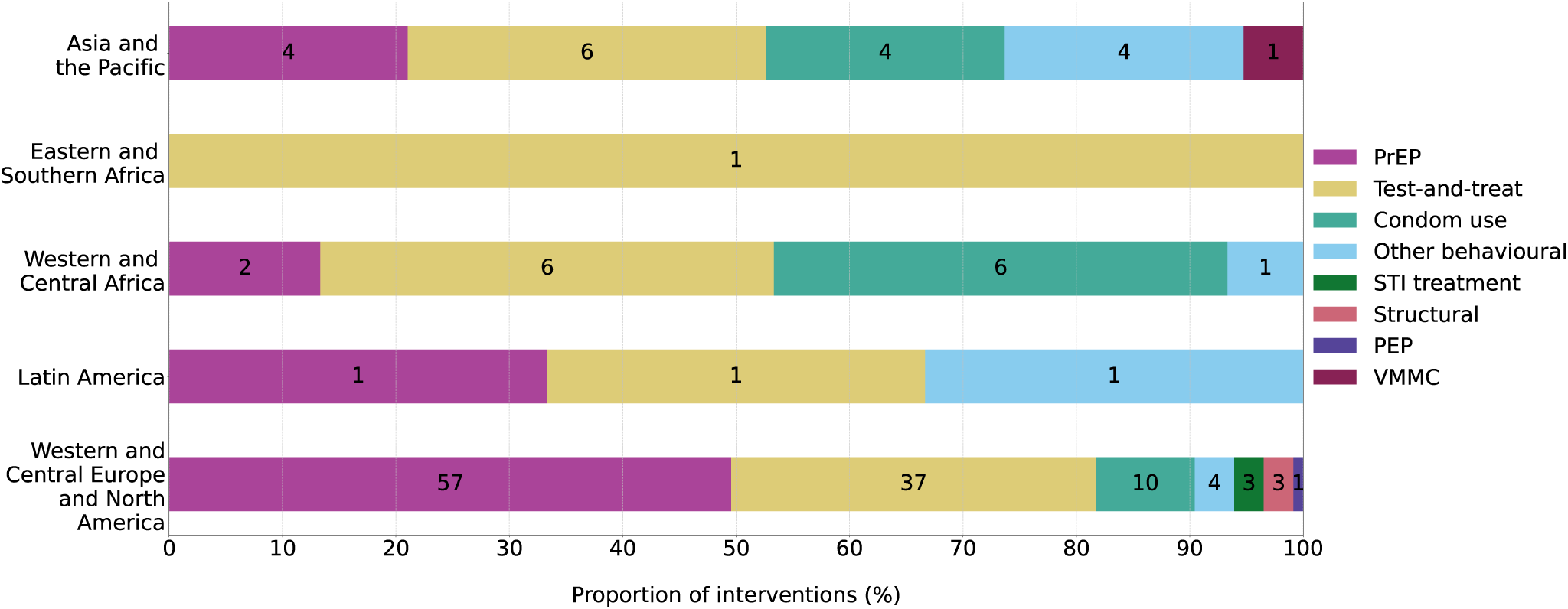
Distribution of modeled interventions by UNAIDS region. The labels on each bar represent the number of times the intervention was included in models. A single study might have included multiple interventions. The study [29] that did not apply to any specific location was not counted. The study [30] that included both the USA and Thailand was counted for two UNAIDS regions. Pre-exposure prophylaxis = PrEP. STI = sexually transmitted infection. PEP = post-exposure prophylaxis. VMMC = voluntary medical male circumcision. The distribution of interventions by UNAIDS region for studies that defined elimination is shown in Figure S2.

### Critical appraisal

Out of the 89 studies investigating the impact of interventions, 39 studies (43.82%) defined criteria for HIV elimination. The critical appraisal results of these 39 studies are summarized in Tables S5 and S6. The model comprehensiveness scores ranged from 0 to 8, although no study received either of these extreme scores. Studies using ABMs scored at least 4, while three studies (15.00%) using DCMs scored 3 or lower [29, 44, 103], and one study (25.00%) using an SCM scored 2 [80]. The average score for ABMs (5.00) was higher than the average scores for DCMs (4.80) and SCMs (4.00). However, three studies (15.00%) using DCMs achieved a score of 7 [28, 71, 86], whereas none of the studies using ABMs did.

Two of the five criteria, reporting outcomes with uncertainty and accounting for adherence to primary interventions, were well satisfied across all studies. All but two studies [80, 105] presented results with some level of uncertainty, whether due to sensitivity analyses, stochastic effects, or multiple parameter sets. Only one study [29] did not account for adherence to the primary interventions. In contrast, the other three criteria were often either not included or only partially included. Sixteen studies (48.72%) (e.g., [28, 71, 85]) attempted to validate their model outputs, although six of these validations were informal or not clearly described [55,56,69,70,81,93]. Fifteen studies (38.46%) (e.g., [48, 80, 93]) modeled open MSM populations, with eight studies using compartmental models and seven using ABMs. Seven studies (17.95%) (e.g., [38, 42, 74]) accounted for sexual risk compensation, more often in compartmental models than in ABMs.

### Elimination definitions

Table 3 provides an overview of the 39 studies that defined HIV elimination. There was no consensus on a single definition of elimination. Ten of the 39 (25.64%) studies used multiple definitions [18,26,42,55,71,81,83,96,101,105]. For these studies, each definition was assessed individually for achievability and feasibility.

**Table 3.**
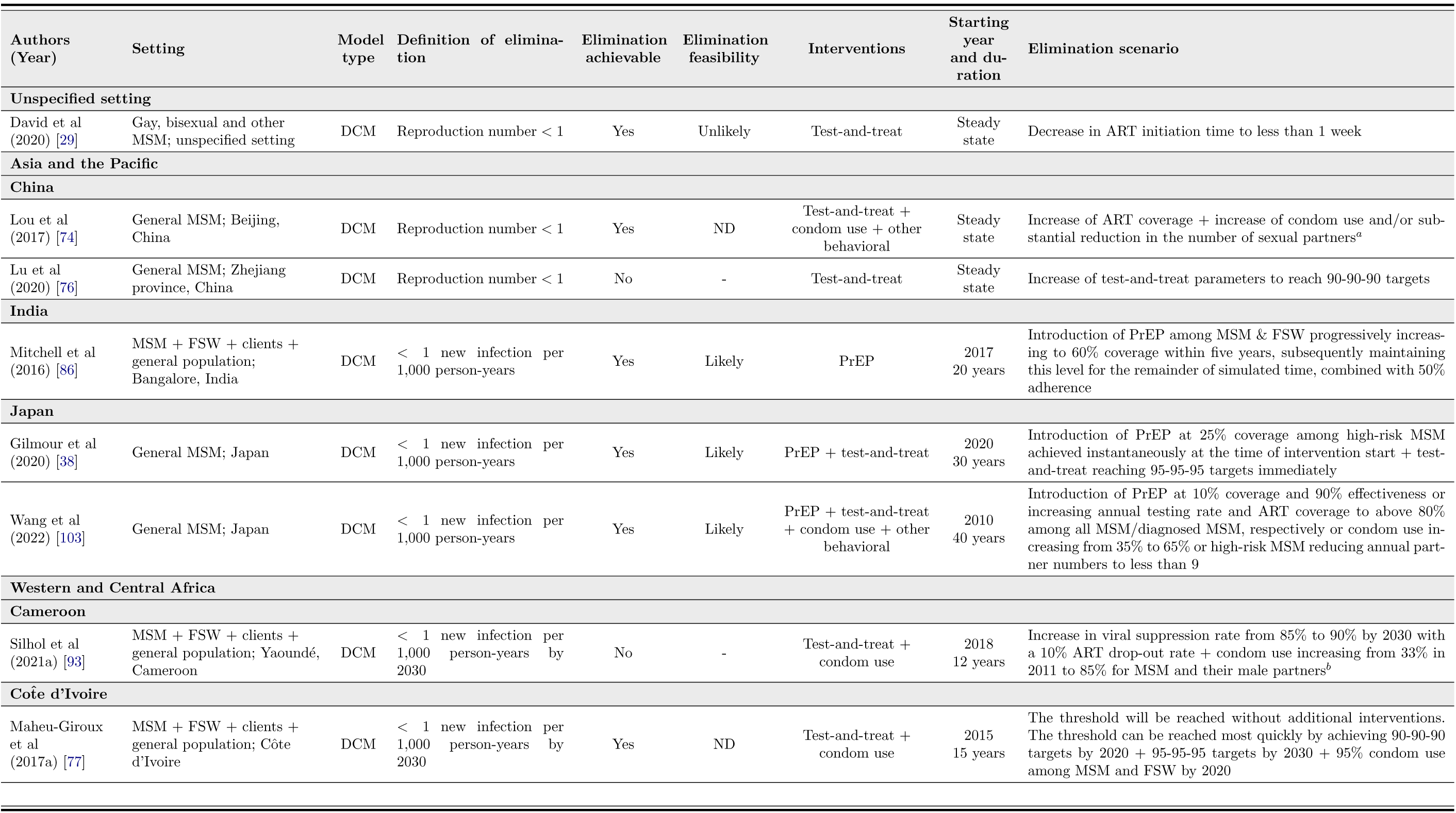
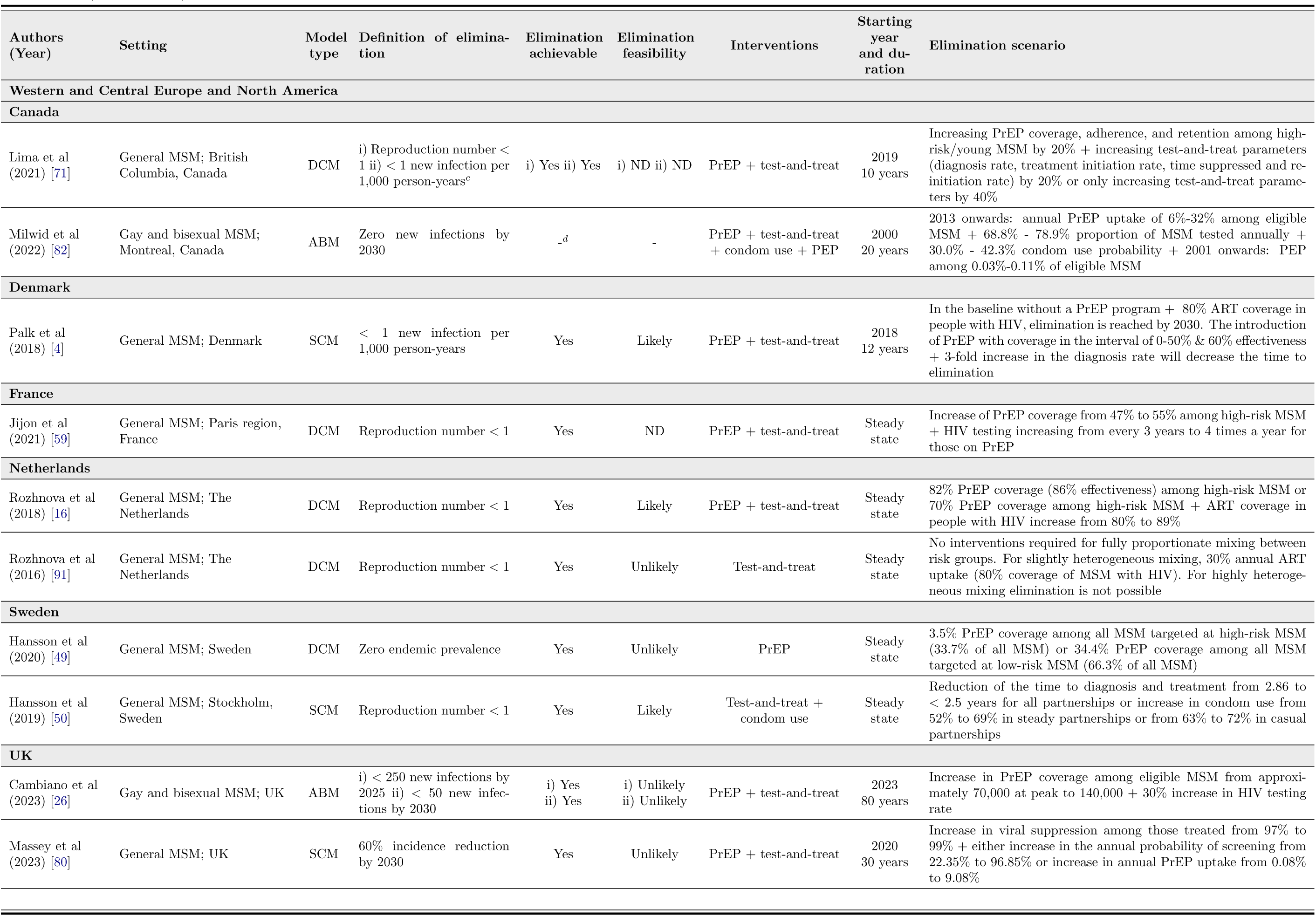
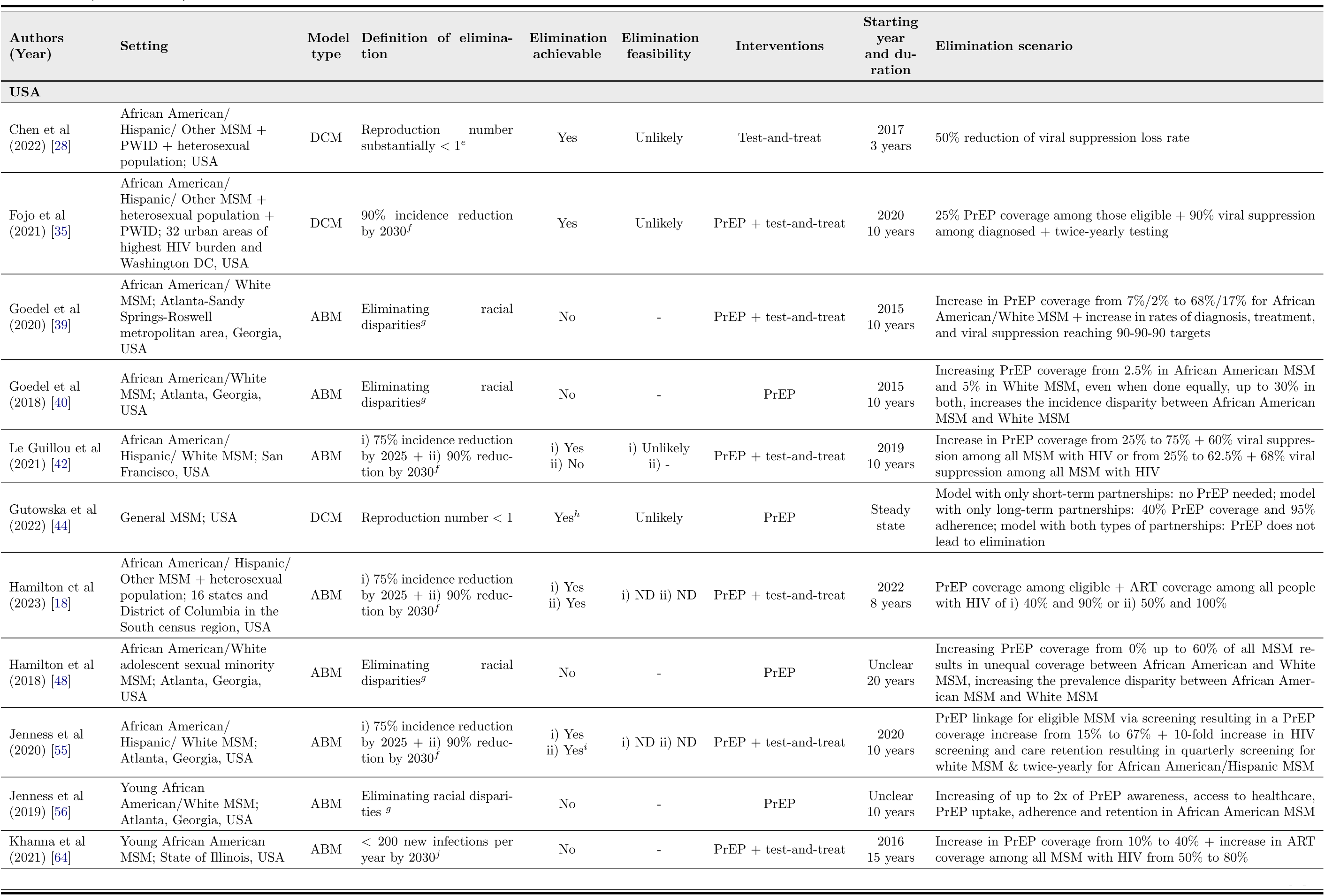
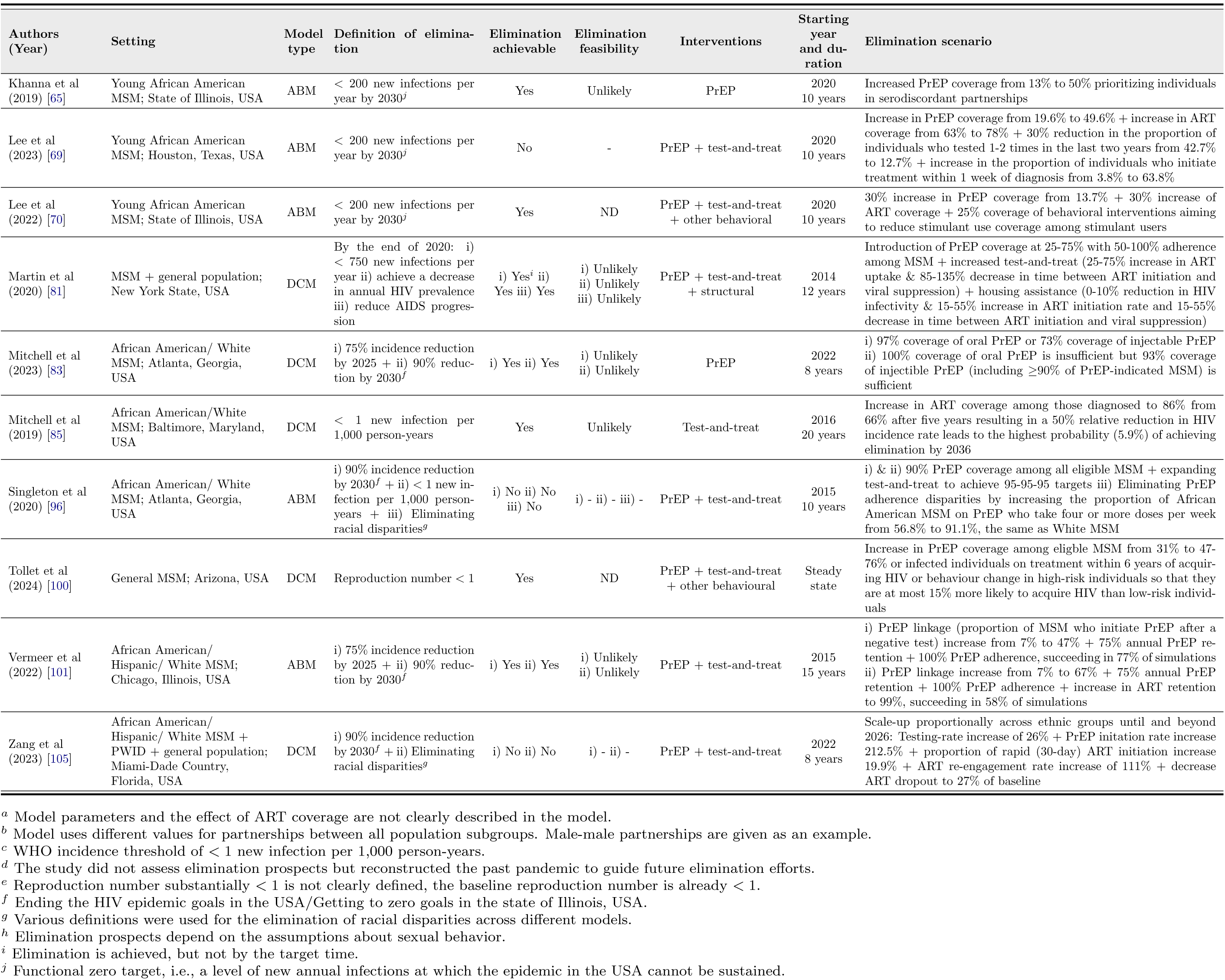
The overview of included studies that defined HIV elimination. In total, 39 out of the 88 studies were included. Elimination achievable indicates whether elimination is technically possible in the modeled scenarios. Elimination feasibility indicates the authors’ judgment on whether it is realistic to implement the modeled elimination scenarios in practice. ND indicates that the feasibility of elimination scenarios was not discussed by the authors. A dash in the elimination feasibility column indicates that a discussion about feasibility is not applicable. It is used for studies where elimination is not possible in the model. The elimination scenario with elimination achievable is the minimum required level of studied interventions. If elimination is not achievable, the elimination scenario describes the level of interventions that came closest to meeting the elimination criteria. MSM = men who have sex with men. TW = transgender women. PWID = people who inject drugs. FSW = female sex workers. SCM = stochastic compartmental model. DCM = deterministic compartmental model. ABM = agent-based model. ART = antiretroviral treatment. PrEP = pre-exposure prophylaxis. ND = not discussed. − = not applicable.

HIV elimination criteria were defined as a threshold for incidence (*n* = 16, 31.37%; e.g., *<* 1 new infection per 1,000 person-years [4, 85, 103]), a percentage reduction in incidence (*n* = 14, 27.45%; e.g., 90% reduction by 2030 from 2020 [35, 42, 83]), reproduction number less than one (*n* = 11, 21.57%; e,g., [29, 44, 76]), elimination of racial disparities in incidence or prevalence (*n* = 6, 11.76% [39, 40, 48, 56, 96, 105]), zero incidence (*n* = 1, 1.96% [82]), zero steady-state prevalence (*n* = 1, 1.96% [49]), and other (*n* = 2, 3.92% [96]).

Elimination definitions varied across model types and UNAIDS regions (Figure 4). A reproduction number less than one was the most frequently used definition in DCMs (*n* = 10, 38.46%) (e.g., [28, 29, 44] and SCMs (*n* = 1, 33.33%) [50], but was absent in ABMs. Conversely, incidence reduction was the most frequent definition in ABMs (*n* = 9, 40.91%) (e.g., [18, 42, 55]) but was seldom used in compartmental models (*n* = 3, 10.34%) [35, 80, 83]. Incidence threshold criteria were used across all model types (*n* = 16, 31.37%) (e.g., [4, 69, 85]), whereas the elimination of racial disparities was almost exclusively studied in ABMs (*n* = 5, 83.33% [39, 40, 48, 56, 96]).

**Figure 4.**
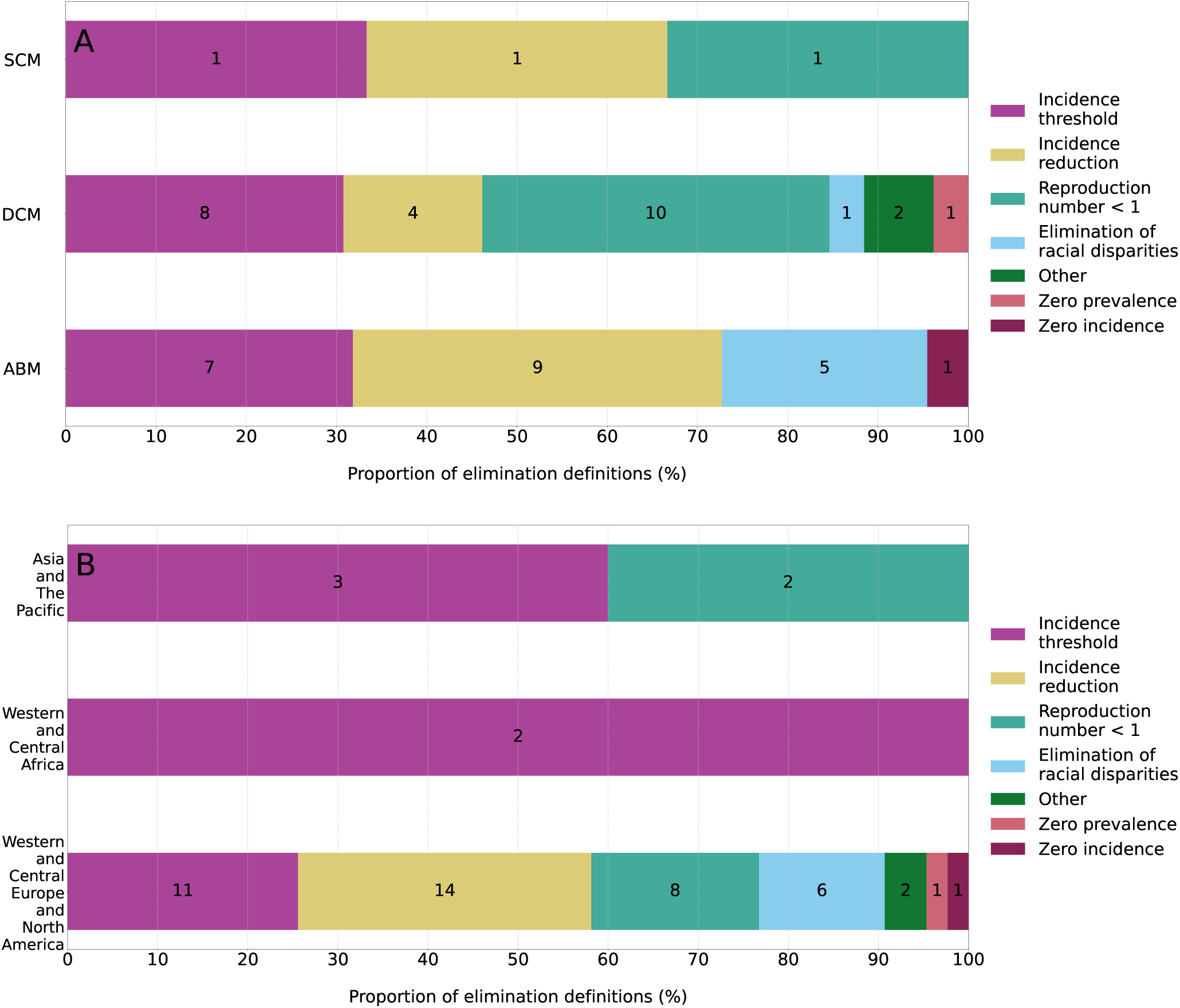
HIV elimination definitions. Distribution of definitions by (**A**) model type and (**B**) UNAIDS region. The labels on each bar represent the number of times the definition was included in studies. A single study might include multiple definitions. In (**B**), the study [29] that did not reference any specific location was not counted. SCM = stochastic compartmental model. DCM = deterministic compartmental model. ABM = agent-based model.

The preferential use of elimination definitions by UNAIDS region (Figure 4 **B**) resulted mainly from (i) ABMs not being used for settings outside Western and Central Europe and North America, specifically the USA, and (ii) studies often aligning their elimination definitions with the national goals of the country where elimination was assessed. For example, incidence reduction accounted for almost half of the definitions used in the USA (*n* = 13, 41.94%) (e.g., [35, 55, 96]), driven by the goals to end the HIV epidemic in the USA, which aim for 75% and 90% reductions in incidence by 2025 and 2030, respectively. In contrast, studies outside of the USA focused on reaching incidence thresholds (e.g., [38, 71, 93]) and reducing the reproduction number below one (e.g., [74, 76, 91]), and did not use incidence reduction as the elimination definition.

### Elimination prospects

In the following, we distinguished between elimination being (i) achievable, if it was technically possible within modeled scenarios, and (ii) feasible, if, based on the authors’ judgment and discussion, the modeled scenarios where elimination was achievable were practical in a real-world context (Table 4). Feasibility was considered (un)likely if the authors judged that the intervention scenario required to achieve elimination in the model was (im)plausible. In 36 out of 50 modeled scenarios (72.00%), elimination was achievable. Using UNAIDS regional stratification, 30 scenarios in Western and Central Europe and North America (e.g., [16,65,85]), one scenario in Western and Central Africa [78], and four scenarios in Asia and the Pacific [38, 74, 86, 103]) could achieve elimination in the model. However, only six (16.67%) of these 36 modeled scenarios were deemed likely to be feasible in practice, half of which were in Western and Central Europe and North America [4, 16, 50] and half in Asia and the Pacific [38, 86, 103]. Eighteen (50.00%) elimination scenarios were deemed unlikely to be feasible in practice, all for settings in Western and Central Europe and North America (e.g., [49, 81, 91]) or without a specific location [29]. The feasibility of twelve (33.33%) scenarios was not discussed by the authors (e.g., [18, 70, 78]).

**Table 4.**
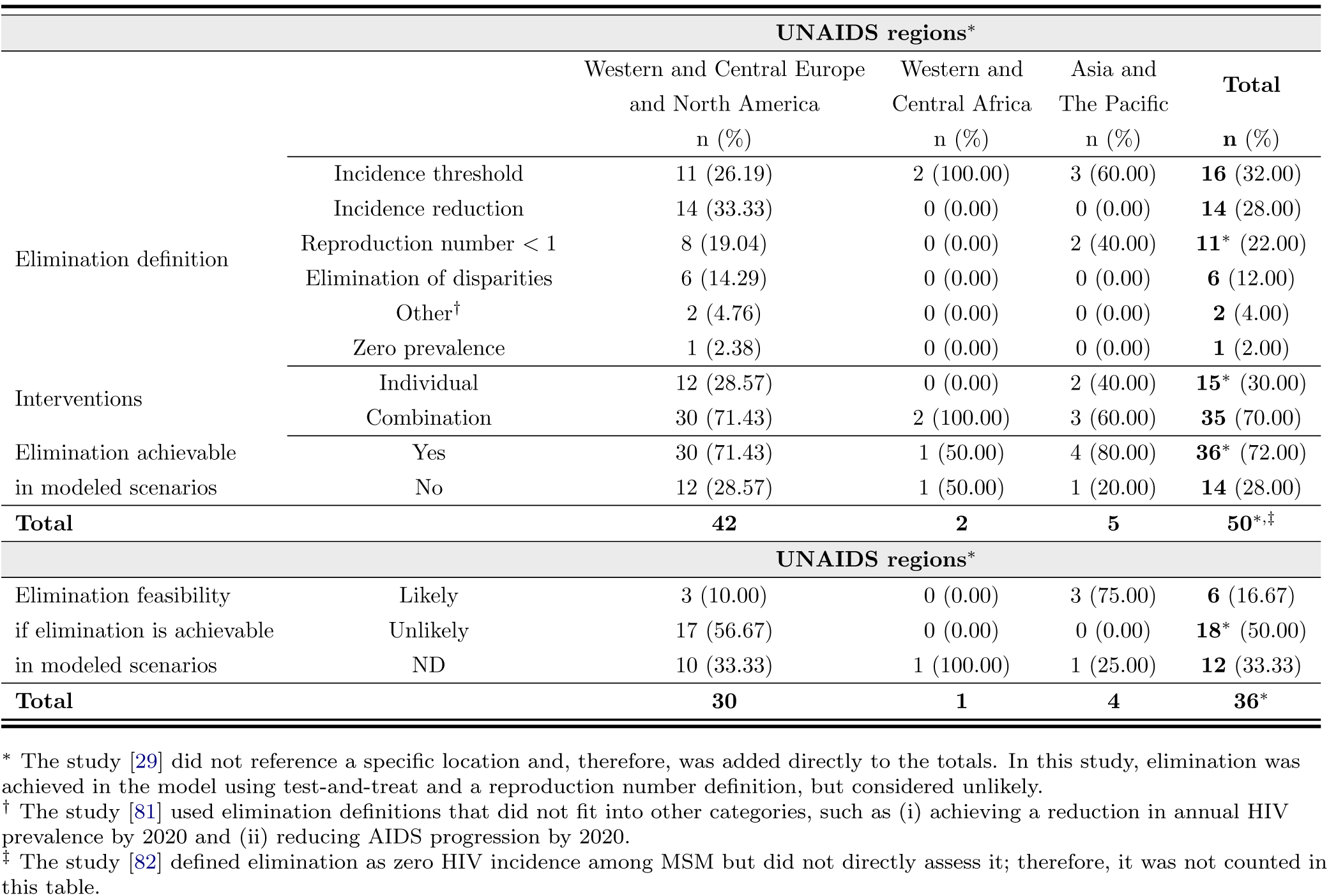
Elimination prospects. MSM = men who have sex with men. SCM = stochastic compartmental model. DCM = deterministic compartmental model. ABM = agent-based model. UNAIDS = The Joint United Nations Programme on HIV/AIDS. ND = not discussed.

Achievability of elimination differed with respect to interventions by UNAIDS region and country (Tables S2 and S3). The majority of scenarios in Western and Central Europe and North America that included PrEP and test-and-treat achieved elimination (*n* = 25, 69.44%, e.g., [4, 44, 59] and *n* = 24, 75.00%, e.g., [18, 42, 71], respectively), while classical behavioral and structural interventions were included less frequently. However, only three elimination scenarios were discussed to be feasible, all for models that consider populations consisting solely of MSM in Europe [4, 16, 50]. Feasible elimination scenarios included an increase of ART coverage and introduction of oral PrEP in the Netherlands [16] and Denmark [4], and a further reduction in time to diagnosis with an increase in condom use in Sweden [50]. In the USA, elimination using a similar composition of interventions was considered unlikely despite the majority of modeled scenarios predicting elimination (e.g., [28, 35, 44]). In Western and Central Africa, test-and-treat and condom use could achieve elimination in half of the modeled scenarios [78], but their feasibility was not discussed. In Asia and the Pacific, modeled elimination scenarios included test-and-treat (*n* = 4, 36.36%), PrEP (*n* = 3, 27.27%), condom use (*n* = 2, 18.18%), and other behavioral (*n* = 2, 18.18%) interventions. Except one scenario that involved test-and-treat only [76], all of them were successful, and their feasibility was also reported as high for India [86] and Japan [38, 103]. The feasible elimination scenarios involved the introduction of oral PrEP and an increase in test-and-treat rates, which could be complemented with condom use and other behavioral interventions. Notably, of the six feasible elimination scenarios, five used a combination of interventions [4, 16, 38, 50, 103], while only one used PrEP as an individual intervention [86]. Elimination prospects stratified by the use of combination and individual interventions are shown in Table S1.

## Discussion

### Underrepresented populations

HIV among MSM is a global problem that transcends geographical borders [1]. To our knowledge, this study is the first to systematically review mathematical modeling studies that assess HIV elimination prospects in this key population worldwide. Our findings show that across all UNAIDS regions, many countries with high HIV burden among MSM were not represented in the recent studies that involve dynamic transmission models. In particular, we did not identify any studies that assess the epidemiological impacts of interventions on MSM in the Caribbean, Eastern Europe and Central Asia, and the Middle East and North Africa. Several factors could contribute to this gap in knowledge, including the lack of high quality sexual behavior and epidemiological data needed for model parameterization, the shortage of local expertise in HIV modeling [107], criminalization of HIV and sodomy, insufficient interest from public health systems in countries where the HIV epidemic in non-MSM populations is more severe than among MSM, poor surveillance, underfunding, discrimination, and stigma.

Notably, a relatively small number of studies targeted MSM in the UNAIDS regions of Western and Central Africa, and Eastern and Southern Africa, collectively known as sub-Saharan Africa [75,77,78,87,92–94,99], where modeling HIV transmission in the general population has traditionally received a lot of attention [108, 109]. This region is known for generalized heterosexual epidemics and a high HIV burden in the general population, but there is also strong evidence of epidemics among MSM [10, 110, 111]. Finally, within Western and Central Europe and North America UNAIDS region, a relatively small number of studies concerned MSM in Europe [4, 16, 26, 49, 50, 59, 67, 80, 89–91] compared to the USA. The likely explanation for this is that modeling methods outside the scope of our review are used to investigate HIV elimination in Europe. For example, back-calculation models have been developed for the Netherlands [6], the UK [5], and Denmark [112] but are not included in our review focused on dynamic transmission models.

### Elimination scenarios

Elimination was achieved in models far more often than authors deemed feasible for real-world implementation (6 out of 36 scenarios). Several reasons could contribute to this discrepancy. Firstly, intervention parameters in models (e.g., PrEP, ART, condom use coverage and adherence, testing rates) can be selected from the maximum possible range, potentially resulting in values that are not achievable in practice (e.g., [42, 81, 83]). Secondly, the feasibility of one-third of modeled elimination scenarios was not discussed by the authors (e.g., [74]), possibly due to the lack of authors with relevant real-world implementation expertise. Thirdly, the six feasible scenarios [4, 16, 38, 50, 86, 103] were obtained in either DCMs or SCMs that were mostly among the least complex models, as described by their comprehensiveness scores. This implies that the results reported by these six models may be overly optimistic.

Our findings show that significant gains in HIV control among MSM have been made in some settings. Elimination is likely in certain Western European countries due to the scale-up of test-and-treat programs, and PrEP emerges as one of the key interventions that can help to reach elimination faster [4]. This aligns with evidence that HIV incidence in countries like the Netherlands and the UK had started to decline with the expansion of test-and-treat programs [5, 6] but declined much further after the introduction of national PrEP programs [113]. Notably, none of the studies in the USA settings considered elimination feasible. This outcome could be partly explained by the use of ABMs and the complexity of the subepidemics in the USA, characterized by strong heterogeneity in transmission among different ethnic and geographical subgroups that require culturally and regionally tailored interventions.

Our findings also highlight inequitable responses in HIV control worldwide. Unlike many studies in Western countries [4, 16, 35, 49, 59, 70, 71, 82, 83] and some studies in Asia [38, 86, 103] that consider PrEP as an essential intervention for faster approach to elimination, studies in Africa still focus on the expansion of treatment and condom use [78, 93]. However, the real-world effectiveness of condoms is undermined by adherence issues. No studies considered elimination scenarios with PrEP introduction among MSM in Africa, potentially due to delays and structural barriers in implementing this intervention [114].

Lastly, our review underscores the importance of combination interventions in increasing the feasibility of elimination. Although the sample of feasible elimination scenarios was small and their conclusions may be overly optimistic, 5 out of 6 considered a combination of interventions [4, 16, 38, 50, 103]. This is consistent with other studies suggesting that a combination of intervention strategies is necessary to control and eliminate HIV [13, 115, 116], and that transmission models should incorporate multiple, simultaneously acting interventions [15].

### Relatedness of elimination definitions

Various definitions of elimination used by different studies reflect their perspectives in terms of modeling and public health. While each elimination definition focuses on a specific aspect of the epidemic dynamics of HIV, these aspects are epidemiologically related. In theory, when the effective reproduction number is below one, the incidence will decrease and eventually elimination will be reached. The smaller the effective reproduction number, the faster the decline in incidence, and the sooner a given incidence threshold will be reached. From a modeling perspective, using the effective reproduction number is attractive (e.g., [50, 59, 103]), because it summarizes the qualitative behavior of the epidemic, and elimination is a consequence of reducing the reproduction number below the threshold of one. From an estimate of the reproduction number, one could, in principle, compute the incidence and the time it takes for the incidence to fall below a threshold. The converse does not hold, i.e., knowing that the incidence is below a certain threshold does not necessarily mean that elimination will be reached in the long term. The incidence could still stabilize at a new lower endemic prevalence if the reproduction number is above one. Therefore, from a modeling perspective, calculating the effective reproduction number has clear advantages over simply calculating incidence. However, an explicit calculation of the reproduction number is only possible for DCMs which explains the frequent use of this definition for these models in our review (e.g., [16,44,71]). For SCMs and ABMs, approximations for a reproduction number can be computed numerically by calculating the average number of secondary cases per infected individual. The involved numerical computations probably explain why none of the ABMs in our review used this definition.

From a public health perspective, it is essential to consider how the path to elimination can be achieved and monitored [117]. This implies that we need elimination definitions based on measurable quantities [118]. Although incidence is not directly observable, it can be estimated from the number of diagnosed cases. More concretely, definitions based on an epidemiological goal such as a threshold (e.g., [4, 64, 85]) or a reduction in incidence (e.g., [18, 26, 35]) can be used in practice to validate model predictions. Both can be measured and monitored over time, and serve as a basis for defining standardized indicators for public health policy evaluation [118]. Therefore, for modeling to contribute effectively to public health policy, it is necessary and valuable to report intervention coverage and incidence, preferably in a format that can easily be compared to standard indicators [117,118]. A clear advantage of using the elimination definition based on an incidence threshold is that incidence can be computed in all model types. Besides offering meaningful guidance to policymakers, this definition would also enhance study comparability within and across different MSM settings.

The elimination of disparities, often discussed in the context of HIV, is a different concept and does not necessarily lead to the elimination of HIV from a population. The goal is to eliminate large differences in HIV incidence between population groups, such as ethnic groups (e.g., [39,40,48]), rather than to eliminate HIV entirely. However, in many epidemiological contexts, it is an important milestone to overall HIV elimination once we consider that groups with high incidence are often characterized by a high risk of HIV acquisition, resulting in effective reproduction numbers in these groups remaining above one the longest. In the final stages before elimination, the groups with the highest reproduction numbers will be the remaining drivers of transmission. Therefore, targeting these groups will be crucial to achieving the final goal of HIV elimination.

### Future elimination modeling

Based on our findings, we identify several areas where HIV elimination modeling in MSM needs to advance. In countries where HIV incidence has dropped considerably in recent years [4–6] and elimination may be possible, modeling should focus not only on interventions that achieve elimination but also on those that sustain it. This conclusion was also supported by [119]. Elimination of transmission among MSM in specific settings may be interpreted as HIV no longer being a public health problem, which could lead to cutbacks in national prevention programs in these communities. For example, scenarios of future changes in the capacity of the national program for oral PrEP are being investigated in the Netherlands [120]. A decrease in PrEP uptake and condom use, combined with a potential increase in sexual risk behavior among MSM [121–123], may lead to a rebound in HIV incidence. A new priority for countries nearing elimination and aiming to sustain it could be interventions targeting HIV infections acquired abroad, either through immigration or travel. Examples of such interventions include offering voluntary HIV testing to incoming migrants or providing PrEP to MSM who travel abroad [4]. Additionally, transmission modeling will need to be complemented with health economic evaluations to guide future interventions that balance the cost of maintaining them and keeping HIV transmission at bay.

Our review highlighted a lack of modeling studies in some regions that have a high HIV prevalence among MSM. Within locations that warrant such attention, a special place is held by so-called island countries. To wit, the Caribbean region, which has the second highest HIV prevalence among MSM across UNAIDS regions [124] and a high volume of migration both within the region and internationally, will require the development of meta-population models. These models including population mobility were absent in our review. Understanding HIV transmission networks and the impact of migration [125] including from Latin America will be crucial for achieving HIV elimination in this region. Meta-population models could also be relevant for MSM populations in island countries in Asia and the Pacific, which have so far received little attention. Movement and migration [125] will generally become increasingly important as they could aid in importing HIV infections from high-prevalence regions to regions nearing elimination, including in the context of Western countries [4].

In our review, almost all studies considered MSM in Africa as part of a wider population consisting of the heterosexual population and key populations other than MSM [77, 78, 87, 92–94, 99]. In contrast to concentrated epidemics among MSM in Western and Central Europe and North America (e,g. [53, 67, 98]), HIV epidemics among MSM and the heterosexual population in Africa are substantially mixed due to men having sex with men and women [126,127]. However, while the numbers of new infections in the overall adult population, sex workers, and their clients in Africa have been falling at the same rate, no such progress has been observed for MSM [3, 10]. If MSM are left behind in the HIV response in Africa, they may continue to be a source of infection for women in the general population in the future, potentially undermining the progress made in eliminating HIV within this population. The inclusion of MSM in transmission models for African countries is needed to inform tailored interventions that ensure equal rates of progress towards HIV elimination for different key populations and the general population [128]. Given the proven benefits of PrEP in Western countries and Asia, exploring its impact on HIV elimination among MSM in Africa is desirable. Additionally, while public health literature highlights the benefits of PEP, the population-level impact of this intervention remains largely unexplored in Africa and globally [129], as does the impact of long-acting injectable PrEP compared to oral PrEP. One study in our review [83] demonstrated that long-acting injectable PrEP programs can achieve HIV elimination at much lower coverage compared to oral PrEP.

Another modeling direction involves understanding the elimination of disparities in HIV burden among diverse subgroups of MSM. As overall incidence decreases, disparities may be exacerbated [128]. Therefore, eliminating disparities is a parallel goal to achieving the overall target for HIV elimination. In our review, this topic has received much attention in the context of epidemics in the USA [39, 40, 48, 56], where pronounced disparities in HIV burden have been observed due to assortative mixing of racial and ethnic subgroups and differences in their interaction with the healthcare system (e.g., testing rates, ART, and PrEP coverage). However, the issue of disparities is not limited to the USA and is relevant to characteristics other than ethnicity in many countries. For example, substantial age differences in HIV prevalence are observed among MSM in Africa [130] and the Caribbean [9]. On the road to elimination, disparities may increase between native and migrant populations [125], as well as between rural and urban communities. In our review, simpler models with fewer stratifications such as DCMs and SCMs predominated outside Western and Central Europe and North America. More complex ABMs will be required to simulate elimination interventions that are sufficiently nuanced to address the vulnerabilities of diverse subgroups of MSM. The need for improved incorporation of characteristics such as ethnicity in future models was also discussed in other literature [15, 108].

Finally, modeling could be useful for understanding how new HIV technologies and biomedical prevention tools might affect transmission dynamics and either strengthen or undermine the feasibility of elimination in different regions. Research efforts are increasingly dedicated to improving the quality of life for people already living with HIV. Biomedical advancements may lead to a potential HIV cure in the future, with a target product profile already formulated [131]. Notably, the target product profile considers the possibility of re-infection and viral rebound after ART-free suppression acceptable. These characteristics imply that introducing an HIV cure may result in new infections among MSM. Investigating future scenarios involving such potential interventions will be necessary to guide their implementation in the context of current HIV elimination efforts.

### Limitations

In the absence of prior systematic reviews on the possibility of global elimination among MSM, we focused our study on mathematical transmission models. These models have the advantage of providing a mechanistic understanding of the transmission process and are frequently used to inform policymakers. The synthesis of elimination definitions and interventions that may lead to elimination in a broader class of models available in the literature (e.g., statistical, back-calculation, decision-analytic, health economic) should be a subject of future research. Additionally, we compared the worldwide location of the studied populations with the HIV prevalence among MSM as reported by the UNAIDS key population Atlas [1]. These UNAIDS data are compiled from public sources and reviewed for quality, though the quality may vary. Other recent estimates of HIV prevalence among MSM cover only specific UNAIDS regions (e.g., [9, 110, 111]) and their use would not alter our overall conclusions about the underrepresentation of MSM populations. However, it is important to note that our decision to exclude studies not published in English may have contributed to the finding that some UNAIDS regions are underrepresented in modeling studies. Furthermore, the presence of various elimination definitions and model types complicated the comparison of efficient intervention scenarios across settings. Different studies modeled similar interventions using a range of modeling paradigms, making cross-comparison challenging. Although we extracted some details on model structures and methodologies, our review primarily focused on elimination definitions and the potential for elimination through various interventions rather than on the structural composition of the models. Therefore, we did not comment on the suitability of the model structure for projecting elimination prospects beyond the criteria included in the comprehensiveness score and used for critical appraisal of studies. In the future, more research should focus on systematic comparative analyses of different models using consistent elimination definitions, epidemiological outcomes. Although it does not specifically define elimination, the study by Eaton et al. [109] serves as an example of such an analysis.

## Conclusion

In conclusion, this systematic review compiled evidence on the possibilities of HIV elimination among MSM world-wide by summarizing findings from mathematical modeling studies. There is a need to intensify modeling efforts to assess elimination prospects among MSM outside Western and Central Europe and North America. Additionally, we analyzed the elimination definitions used in current studies and recommended new research directions for modeling to support a coordinated global response to HIV elimination among MSM.

## Contributors

G.R. supervised the study. G.R. and J.A.R. wrote the manuscript. J.A.R., A.T., and G.R. extracted data. J.A.R. analyzed data. J.A.R. and A.T. performed the critical appraisal of the studies. M.E.K., J.H.H.M.v.d.W., and A.T. made substantial contributions to the discussions and commented on the manuscript.

## Data sharing

The extracted data are available in Appendix 2.

## Data Availability

All data are avaiable in appendix 2. (Not available in preprint publication).

## Acknowledgments

GR and AT were supported by the Aidsfonds Netherlands (grant number P-53902). GR and JAR were supported by the VERDI project (101045989), funded by the European Union. Views and opinions expressed are those of the author(s) only and do not necessarily reflect those of the European Union or the Health and Digital Executive Agency. Neither the European Union nor the granting authority can be held responsible for them. We thank Kevin Jenniskens (University Medical Center Utrecht and Cochrane Netherlands), Marleen Werkman (University Medical Center Utrecht), Noor Godijk van Merkestein (GGD Limburg-Noord), Maria Xiridou (The National Institute for Public Health and The Environment), and members of the Infectious Disease Modeling Group (University Medical Center Utrecht) for useful discussions.

## Supplementary material

The full PRISMA checklist is available in Appendix 1. Additional results are shown in Figures S1-S2 and Tables S1-S6.

## Correspondence

Correspondence and material requests should be addressed to Dr. Ganna Rozhnova, Julius Center for Health Sciences and Primary Care, University Medical Center Utrecht, P.O. Box 85500 Utrecht, The Netherlands.

## References

[1] The Joint United Nations Programme on HIV/AIDS (UNAIDS). UNAIDS Key Population Atlas; 2024. Available from: https://kpatlas.unaids.org/dashboard.

[2] The Joint United Nations Programme on HIV/AIDS (UNAIDS). Global HIV & AIDS statistics — Fact sheet; 2024. Available from: https://www.unaids.org/sites/default/files/media_asset/UNAIDS_FactSheet_en.pdf.

[3] Korenromp EL, Sabin K, Stover J, Brown T, Johnson LF, Martin-Hughes R, et al. New HIV infections among key populations and their partners in 2010 and 2022, by world region: a multisources estimation. Journal of acquired immune deficiency syndromes. 2024;95(1):S34–S45. doi:10.1097/QAI.0000000000003340.

[4] Palk L, Gerstoft J, Obel N, Blower S. A modeling study of the Danish HIV epidemic in men who have sex with men: travel, pre-exposure prophylaxis and elimination. Scientific reports. 2018;8(1):16003. doi:10.1038/s41598-018-33570-0.

[5] Brizzi F, Birrell PJ, Kirwan P, Ogaz D, Brown AE, Delpech VC, et al. Tracking elimination of HIV transmission in men who have sex with men in England: a modelling study. The Lancet HIV. 2021;8(7):e440–e448. doi:10.1016/S2352-3018(21)00044-8.

[6] van Sighem A, Wit F, Boyd A, Smit C, Jongen V, Koole J. Monitoring Report 2023. Human Immunodeficiency Virus (HIV) Infection in the Netherlands. Amsterdam: Stichting hiv monitoring; 2023. Available from: https://www.hiv-monitoring.nl.

[7] Mumtaz GR, Chemaitelly H, Abu-Raddad LJ. In: Laher I, editor. The HIV Epidemic in the Middle East and North Africa: Key Lessons. Cham: Springer International Publishing; 2021. p. 3053–3079. Available from: 10.1007/978-3-030-36811-1_139.

[8] Mumtaz GR, Chemaitelly H, AlMukdad S, Osman A, Fahme S, Rizk NA, et al. Status of the HIV epidemic in key populations in the Middle East and north Africa: knowns and unknowns. The Lancet HIV. 2022;9(7):e506– e516. doi:10.1016/S2352-3018(22)00093-5.

[9] Coelho LE, Torres TS, Veloso VG, Grinsztejn B, Jalil EM, Wilson EC, et al. The prevalence of HIV among men who have sex with men (MSM) and young MSM in Latin America and the Caribbean: a systematic review. AIDS and behavior. 2021;25(10):3223–3237. doi:10.1007/s10461-021-03180-5.

[10] Stannah J, Soni N, Lam JKS, Giguère K, Mitchell KM, Kronfli N, et al. Trends in HIV testing, the treatment cascade, and HIV incidence among men who have sex with men in Africa: a systematic review and meta-analysis. The Lancet HIV. 2023;10(8):e528–e542. doi:10.1016/S2352-3018(23)00111-X.

[11] McCormack S, Dunn DT, Desai M, Dolling DI, Gafos M, Gilson R, et al. Pre-exposure prophylaxis to prevent the acquisition of HIV-1 infection (PROUD): effectiveness results from the pilot phase of a pragmatic open-label randomised trial. The Lancet. 2016;387(10013):53–60. doi:10.1016/S0140-6736(15)00056-2.

[12] Granich RM, Gilks CF, Dye C, De Cock KM, Williams BG. Universal voluntary HIV testing with immediate antiretroviral therapy as a strategy for elimination of HIV transmission: a mathematical model. Lancet. 2008;373(9657):48–57. doi:10.1016/S0140-6736(08)61697-9.

[13] van Sighem A, van der Valk M. Moving towards zero new HIV infections: The importance of combination prevention. The Lancet regional health – Western Pacific. 2022;25. doi:10.1016/j.lanwpc.2022.100558.

[14] Garnett GP. An introduction to mathematical models in sexually transmitted disease epidemiology. Sex Transm Infect. 2002;78(1):7–12. doi:10.1136/sti.78.1.7.

[15] Giddings R, Indravudh P, Medley GF, Bozzani F, Gafos M, Malhotra S, et al. Infectious disease modelling of HIV prevention interventions: a systematic review and narrative synthesis of compartmental models. PharmacoEconomics. 2023;41(6):693–707. doi:10.1007/s40273-023-01260-z.

[16] Rozhnova G, Heijne J, Bezemer D, van Sighem A, Presanis A, De Angelis D, et al. Elimination prospects of the Dutch HIV epidemic among men who have sex with men in the era of preexposure prophylaxis. AIDS. 2018;32(17):2615–2623. doi:10.1097/QAD.0000000000002050.

[17] The Joint United Nations Programme on HIV/AIDS (UNAIDS). Understanding fast-track accelerating action to end the AIDS epidemic by 2030; 2015. Available from: https://www.unaids.org/sites/default/files/media_asset/201506_JC2743_Understanding_FastTrack_en.pdf.

[18] Hamilton DT, Hoover KW, Smith DK, Delaney KP, Wang LY, Li J, et al. Achieving the “Ending the HIV Epidemic in the U.S.” incidence reduction goals among at-risk populations in the South. BMC public health. 2023;23(1):716. doi:10.1186/s12889-023-15563-5.

[19] Page MJ, McKenzie JE, Bossuyt PM, Boutron I, Hoffmann TC, Mulrow CD, et al. The PRISMA 2020 statement: an updated guideline for reporting systematic reviews. BMJ. 2021;372:n71. doi:10.1136/bmj.n71.

[20] World Health Organization. Consolidated guidelines on the use of antiretroviral drugs for treating and preventing HIV infection: recommendations for a public health approach, 2nd ed; r. Available from: https://www.who.int/publications/i/item/9789241549684.

[21] Balasubramanian R, Kasaie P, Schnure M, Dowdy DW, Shah M, Fojo AT. Projected impact of expanded long-acting injectable PrEP use among men who have sex with men on local HIV epidemics. Journal of acquired immune deficiency syndromes. 2022;91(2):144–150. doi:10.1097/QAI.0000000000003029.

[22] Booton RD, Ong JJ, Lee A, Liu A, Huang W, Wei C, et al. Modelling the impact of an HIV testing intervention on HIV transmission among men who have sex with men in China. HIV medicine. 2021;22(6):467–477. doi:10.1111/hiv.13063.

[23] Bórquez A, Rich K, Farrell M, Degenhardt L, McKetin R, Tran LT, et al. Integrating HIV pre-exposure prophylaxis and harm reduction among men who have sex with men and transgender women to address intersecting harms associated with stimulant use: a modelling study. Journal of the international AIDS society. 2020;23 Suppl 1(Suppl 1):e25495. doi:10.1002/jia2.25495.

[24] Buchanan AL, Park CJ, Bessey S, Goedel WC, Murray EJ, Friedman SR, et al. Spillover benefit of pre-exposure prophylaxis for HIV prevention: evaluating the importance of effect modification using an agent-based model. Epidemiology and infection. 2022;150:e192. doi:10.1017/S0950268822001650.

[25] Buchanan AL, Bessey S, Goedel WC, King M, Murray EJ, Friedman SR, et al. Disseminated effects in agent-based models: A potential outcomes framework and application to inform preexposure prophylaxis coverage levels for HIV prevention. American journal of epidemiology. 2021;190(5):939–948. doi:10.1093/aje/kwaa239.

[26] Cambiano V, Miners A, Lampe FC, McCormack S, Gill ON, Hart G, et al. The effect of combination prevention strategies on HIV incidence among gay and bisexual men who have sex with men in the UK: a model-based analysis. Lancet HIV. 2023;10(11):e713–e722. doi:10.1016/S2362-3018(23)00204-7.

[27] Chan PA, Goedel WC, Nunn AS, Sowemimo-Coker G, Galárraga O, Prosperi M, et al. Potential impact of interventions to enhance retention in care during real-world HIV pre-exposure prophylaxis implementation. AIDS patient care STDS. 2019;33(10):434–439. doi:10.1089/apc.2019.0064.

[28] Chen YH, Farnham PG, Hicks KA, Sansom SL. Estimating the HIV effective reproduction number in the United States and evaluating HIV elimination strategies. Journal of public health management and practice. 2022;28(2):152–161. doi:10.1097/PHH.0000000000001397.

[29] David JF, Lima VD, Zhu J, Brauer F. A co-interaction model of HIV and syphilis infection among gay, bisexual and other men who have sex with men. Infectious disease modelling. 2020;5:855–870. doi:10.1016/j.idm.2020.10.008.

[30] Dimitrov D, Moore JR, Wood D, Mitchell KM, Li M, Hughes JP, et al. Predicted effectiveness of daily and nondaily preexposure prophylaxis for men who have sex with men based on sex and pill-taking patterns from the human immuno virus prevention trials network 067/ADAPT study. Clinical infectious diseases. 2019;71(2):249–255. doi:10.1093/cid/ciz799.

[31] Dimitrov D, Wood D, Ulrich A, Swan DA, Adamson B, Lama JR, et al. Projected effectiveness of HIV detection during early infection and rapid ART initiation among MSM and transgender women in Peru: a modeling study. Infectious disease modelling. 2019;4:73–82. doi:10.1016/j.idm.2019.04.001.

[32] Doyle CM, Milwid RM, Cox J, Xia Y, Lambert G, Tremblay C, et al. Population-level effectiveness of pre-exposure prophylaxis for HIV prevention among men who have sex with men in Montreal (Canada): a modelling study of surveillance and survey data. Journal of the international AIDS society. 2023;26(12):e26194. doi:10.1002/jia2.26194.

[33] Drabo EF, Moucheraud C, Nguyen A, Garland WH, Holloway IW, Leibowitz A, et al. Using microsimulation modeling to inform EHE implementation strategies in Los Angeles County. Journal of acquired immune deficiency syndromes. 2022;90(S1):S167–S176. doi:10.1097/QAI.0000000000002977.

[34] Elion RA, Kabiri M, Mayer KH, Wohl DA, Cohen J, Beaubrun AC, et al. Estimated impact of targeted pre-exposure prophylaxis: strategies for men who have sex with men in the United States. International Journal of Environmental Research and Public Health. 2019;16(9). doi:10.3390/ijerph16091592.

[35] Fojo AT, Schnure M, Kasaie P, Dowdy DW, Shah M. What will it take to end HIV in the United States? : A comprehensive, local-level modeling study. Annals of internal medicine. 2021;174(11):1542–1553. doi:10.7326/M21-1501.

[36] Fraser H, Borquez A, Stone J, Abramovitz D, Brouwer KC, Goodman-Meza D, et al. Overlapping key populations and HIV transmission in Tijuana, Mexico: a modelling analysis of epidemic drivers. AIDS and behavior. 2021;25(11):3814–3827. doi:10.1007/s10461-021-03361-2.

[37] Gantenberg JR, King M, Montgomery MC, Galárraga O, Prosperi M, Chan PA, et al. Improving the impact of HIV pre-exposure prophylaxis implementation in small urban centers among men who have sex with men: an agent-based modelling study. PLoS One. 2018;13(7):e0199915. doi:10.1371/journal.pone.0199915.

[38] Gilmour S, Peng L, Li J, Oka S, Tanuma J. New strategies for prevention of HIV among Japanese men who have sex with men: a mathematical model. Scientific Reports. 2020;10(1):18187. doi:10.1038/s41598-020-75182-7.

[39] Goedel WC, Bessey S, Lurie MN, Biello KB, Sullivan PS, Nunn AS, et al. Projecting the impact of equity-based preexposure prophylaxis implementation on racial disparities in HIV incidence among MSM. AIDS. 2020;34(10):1509–1517. doi:10.1097/QAD.0000000000002577.

[40] Goedel WC, King MRF, Lurie MN, Nunn AS, Chan PA, Marshall BDL. Effect of racial inequities in pre-exposure prophylaxis use on racial disparities in HIV incidence among men who have sex with men: a modeling study. Journal of acquired immune deficiency syndromes. 2018;79(3):323–329. doi:10.1097/QAI.0000000000001817.

[41] Goodreau SM, Hamilton DT, Jenness SM, Sullivan PS, Valencia RK, Wang LY, et al. Targeting human immunodeficiency virus pre-exposure prophylaxis to adolescent sexual minority males in higher prevalence areas of the United States: a modeling study. Journal of adolescent health. 2018;62(3):311–319. doi:10.1016/j.jadohealth.2017.09.023.

[42] Le Guillou A, Buchbinder S, Scott H, Liu A, Havlir D, Scheer S, et al. Population impact and efficiency of improvements to HIV PrEP under conditions of high ART coverage among San Francisco men who have sex with men. Journal of acquired immune deficiency syndromes. 2021;88(4):340–347. doi:10.1097/QAI.0000000000002781.

[43] Gurski K, Hoffman K. Staged HIV transmission and treatment in a dynamic model with long-term partnerships. Journal of mathematical biology. 2023;86(5):74. doi:10.1007/s00285-023-01885-w.

[44] Gutowska SJ, Hoffman KA, Gurski KF. The effect of PrEP uptake and adherence on the spread of HIV in the presence of casual and long-term partnerships. Mathematical Biosciences and Engineering. 2022;19(12):11903– 11934. doi:10.3934/mbe.2022555.

[45] Hamilton DT, Rosenberg ES, Sullivan PS, Wang LY, Dunville RL, Barrios LC, et al. Modeling the impact of PrEP programs for adolescent sexual minority males based on empirical estimates for the PrEP continuum of care. Journal of adolescent health. 2020;68(3):488–496. doi:10.1016/j.jadohealth.2020.06.041.

[46] Hamilton DT, Katz DA, Luo W, Stekler JD, Rosenberg ES, Sullivan PS, et al. Effective strategies to promote HIV self-testing for men who have sex with men: evidence from a mathematical model. Epidemics. 2021;37:100518. doi:10.1016/j.epidem.2021.100518.

[47] Hamilton DT, Rosenberg ES, Jenness SM, Sullivan PS, Wang LY, Dunville RL, et al. Modeling the joint effects of adolescent and adult PrEP for sexual minority males in the United States. PLoS One. 2019;14(5):e0217315. doi:10.1371/journal.pone.0217315.

[48] Hamilton DT, Goodreau SM, Jenness SM, Sullivan PS, Wang LY, Dunville RL, et al. Potential impact of HIV preexposure prophylaxis among black and white adolescent sexual minority males. American journal of public health. 2018;108(S4):S284–S291. doi:10.2105/AJPH.2018.304471.

[49] Hansson D, Strömdahl S, Leung KY, Britton T. Introducing pre-exposure prophylaxis to prevent HIV acquisition among men who have sex with men in Sweden: insights from a mathematical pair formation model. BMJ Open. 2020;10(2):e033852. doi:10.1136/bmjopen-2019-033852.

[50] Hansson D, Leung KY, Britton T, Strömdahl S. A dynamic network model to disentangle the roles of steady and casual partners for HIV transmission among MSM. Epidemics. 2019;27:66–76. doi:10.1016/j.epidem.2019.02.001.

[51] Hotton AL, Lee F, Sheeler D, Ozik J, Collier N, Edali M, et al. Impact of post-incarceration care engagement interventions on HIV transmission among young Black men who have sex with men and their sexual partners: an agent-based network modeling study. The Lancet regional health - Americas. 2023;28:100628. doi:10.1016/j.lana.2023.100628.

[52] Irvine MA, Salway T, Grennan T, Wong J, Gilbert M, Coombs D. Predicting the impact of clustered risk and testing behaviour patterns on the population-level effectiveness of pre-exposure prophylaxis against HIV among gay, bisexual and other men who have sex with men in Greater Vancouver, Canada. Epidemics. 2019;30:100360. doi:10.1016/j.epidem.2019.100360.

[53] Irvine MA, Konrad BP, Michelow W, Balshaw R, Gilbert M, Coombs D. A novel Bayesian approach to predicting reductions in HIV incidence following increased testing interventions among gay, bisexual and other men who have sex with men in Vancouver, Canada. Journal of the royal society interface. 2018;15(140). doi:10.1098/rsif.2017.0849.

[54] Jenness SM, Le Guillou A, Chandra C, Mann LM, Sanchez T, Westreich D, et al. Projected HIV and bacterial sexually transmitted infection incidence following COVID-19-related sexual distancing and clinical service interruption. Journal of infectious diseases. 2021;223(6):1019–1028. doi:10.1093/infdis/jiab051.

[55] Jenness SM, Johnson JA, Hoover KW, Smith DK, Delaney KP. Modeling an integrated HIV prevention and care continuum to achieve the ending the HIV epidemic goals. AIDS. 2020;34(14):2103–2113. doi:10.1097/QAD.0000000000002681.

[56] Jenness SM, Maloney KM, Smith DK, Hoover KW, Goodreau SM, Rosenberg ES, et al. Addressing gaps in HIV preexposure prophylaxis care to reduce racial disparities in HIV incidence in the United States. American journal of epidemiology. 2019;188(4):743–752. doi:10.1093/aje/kwy230.

[57] Jenness SM, Sharma A, Goodreau SM, Rosenberg ES, Weiss KM, Hoover KW, et al. Individual HIV risk versus population impact of risk compensation after HIV preexposure prophylaxis initiation among men Who have sex with men. PLoS One. 2017;12(1):e0169484. doi:10.1371/journal.pone.0169484.

[58] Jenness SM, Goodreau SM, Rosenberg E, Beylerian EN, Hoover KW, Smith DK, et al. Impact of the Centers for Disease Control’s HIV preexposure prophylaxis guidelines for men who have sex with men in the United States. Journal of infectious diseases. 2016;214(12):1800–1807. doi:10.1093/infdis/jiw223.

[59] Jijón S, Molina JM, Costagliola D, Supervie V, Breban R. Can HIV epidemics among MSM be eliminated through participation in preexposure prophylaxis rollouts? AIDS. 2021;35(14):2347–2354. doi:10.1097/QAD.0000000000003012.

[60] Jones J, Le Guillou A, Gift TL, Chesson H, Bernstein KT, Delaney KP, et al. Effect of screening and treatment for gonorrhea and chlamydia on HIV incidence among men who have sex with men in the United States: a modeling analysis. Sexually Transmitted Diseases. 2022;49(10). doi:10.1097/OLQ.0000000000001685.

[61] Kasaie P, Schumacher CM, Jennings JM, Berry SA, Tuddenham SA, Shah MS, et al. Gonorrhoea and chlamydia diagnosis as an entry point for HIV pre-exposure prophylaxis: a modelling study. BMJ open. 2019;9(3). doi:10.1136/bmjopen-2018-023453.

[62] Kasaie P, Berry SA, Shah MS, Rosenberg ES, Hoover KW, Gift TL, et al. Impact of providing preexposure prophylaxis for human immunodeficiency virus at clinics for sexually transmitted infections in Baltimore City: an agent-based model. Sexually transmitted diseases. 2018;45(12):791–797. doi:10.1097/OLQ.0000000000000882.

[63] Kasaie P, Pennington J, Shah MS, Berry SA, German D, Flynn CP, et al. The impact of preexposure prophylaxis among men who have sex with men: an individual-based model. Journal of acquired immune deficiency syndromes. 2017;75(2):175–183. doi:10.1097/QAI.0000000000001354.

[64] Khanna AS, Edali M, Ozik J, Collier N, Hotton A, Skwara A, et al. Projecting the number of new HIV infections to formulate the “Getting to Zero” strategy in Illinois, USA. Mathematical Biosciences and Engineering. 2021;18(4):3922–3938. doi:10.3934/mbe.2021196.

[65] Khanna AS, Schneider JA, Collier N, Ozik J, Issema R, di Paola A, et al. A modeling framework to inform preexposure prophylaxis initiation and retention scale-up in the context of ’Getting to Zero’ initiatives. AIDS. 2019;33(12):1911–1922. doi:10.1097/QAD.0000000000002290.

[66] Khurana N, Yaylali E, Farnham PG, Hicks KA, Allaire BT, Jacobson E, et al. Impact of improved HIV care and treatment on PrEP effectiveness in the United States, 2016–2020. Journal of acquired immune deficiency syndromes. 2018;78(4):399–405. doi:10.1097/QAI.0000000000001707.

[67] Kusejko K, Marzel A, Hampel B, Bachmann N, Nguyen H, Fehr J, et al. Quantifying the drivers of HIV transmission and prevention in men who have sex with men: a population model-based analysis in Switzerland. HIV medicine. 2018;19(10):688–697. doi:10.1111/hiv.12660.

[68] Labs J, Nunn AS, Chan PA, Bessey S, Park CJ, Marshall BDL, et al. Projected effects of disruptions to human immunodeficiency virus (HIV) prevention services during the Coronavirus Disease 2019 pandemic among Black/African American men who have sex with men in an ending the HIV epidemic priority jurisdiction. Open forum infectious diseases. 2022;9(7):ofac274. doi:10.1093/ofid/ofac274.

[69] Lee F, Khanna AS, Hallmark CJ, Lavingia R, McNeese M, Zhao J, et al. Expanding medicaid to reduce human immunodeficiency virus transmission in Houston, Texas: insights from a modeling study. Medical care. 2023;61(1):12–19. doi:10.1097/MLR.0000000000001772.

[70] Lee F, Sheeler D, Hotton A, Vecchio ND, Flores R, Fujimoto K, et al. Stimulant use interventions may strengthen ‘Getting to Zero’ HIV elimination initiatives in Illinois: insights from a modeling study. International journal of drug policy. 2022;103:103628. doi:10.1016/j.drugpo.2022.103628.

[71] Lima VD, Zhu J, Card KG, Lachowsky NJ, Chowell-Puente G, Wu Z, et al. Can the combination of TasP and PrEP eliminate HIV among MSM in British Columbia, Canada? Epidemics. 2021;35:100461. 10.1016/j.epidem.2021.100461.

[72] LeVasseur MT, Goldstein ND, Tabb LP, Olivieri-Mui BL, Welles SL. The effect of PrEP on HIV incidence among men who have sex with men in the context of condom use, treatment as prevention, and seroadaptive practices. Journal of acquired immune deficiency syndromes. 2018;77(1):31–40. doi:10.1097/QAI.0000000000001555.

[73] Lou J, Cheng J, Li Y, Zhang C, Xing H, Ruan Y, et al. Comparison of different strategies for controlling HIV/AIDS spreading in MSM. Infectious disease modelling. 2018;3:293–300. doi:10.1016/j.idm.2018.10.002.

[74] Lou J, Hu P, Qian HZ, Ruan Y, Jin Z, Xing H, et al. Expanded antiretroviral treatment, sexual networks, and condom use: treatment as prevention unlikely to succeed without partner reduction among men who have sex with men in China. PLOS ONE. 2017;12(4):1–18. doi:10.1371/journal.pone.0171295.

[75] Lyons CE, Stokes-Cawley OJ, Simkin A, Bowring AL, Mfochive Njindam I, Njoya O, et al. Modeling the potential impact of pre-exposure prophylaxis for HIV among men who have sex with men in Cameroon. BMC infectious diseases. 2022;22(1):751. doi:10.1186/s12879-022-07738-z.

[76] Lu Z, Wang L, Wang P, Xing H, Fu G, Yan H, et al. A mathematical model for HIV prevention and control among men who have sex with men in China. Epidemiology and infection. 2020;148(224):1–9. doi:10.1017/S0950268820000850.

[77] Maheu-Giroux M, Vesga JF, Diabaté S, Alary M, Baral S, Diouf D, et al. Changing dynamics of HIV transmission in Ĉote d’Ivoire: modeling who acquired and transmitted infections and estimating the impact of past HIV interventions (1976–2015). Journal of acquired immune deficiency syndromes. 2017;75(5):517–527. doi:10.1097/QAI.0000000000001434.

[78] Maheu-Giroux M, Vesga JF, Diabaté S, Alary M, Baral S, Diouf D, et al. Population-level impact of an accelerated HIV response plan to reach the UNAIDS 90-90-90 target in Ĉote d’Ivoire: insights from mathematical modeling. PLOS medicine. 2017;14(6):1–18. doi:10.1371/journal.pmed.1002321.

[79] Marshall BDL, Goedel WC, King MRF, Singleton A, Durham DP, Chan PA, et al. Potential effectiveness of long-acting injectable pre-exposure prophylaxis for HIV prevention in men who have sex with men: a modelling study. Lancet HIV. 2018;5(9):e498–e505. doi:10.1016/S2352-3018(18)30097-3.

[80] Massey K, Vardanega V, Chaponda M, Eddowes LA, Hearmon N. Investigating zero transmission of HIV in the MSM population: a UK modelling case study. Archives of public health. 2023;81(1):201. doi:10.1186/s13690-023-01178-0.

[81] Martin EG, MacDonald RH, Gordon DE, Swain CA, O’Donnell T, Helmeset J, et al. Simulating the end of AIDS in New York: using participatory dynamic modeling to improve implementation of the ending the epidemic initiative. Public health reports. 2020;135(1 suppl):158S–171S. doi:10.1177/0033354920935069.

[82] Milwid RM, Xia Y, Doyle CM, Cox J, Lambert G, Thomas R, et al. Past dynamics of HIV transmission among men who have sex with men in Montŕeal, Canada: a mathematical modeling study. BMC infectious diseases. 2022;22(1):233. doi:10.1186/s12879-022-07207-7.

[83] Mitchell KM, Boily MC, Hanscom B, Moore M, Todd J, Paz-Bailey G, et al. Estimating the impact of HIV PrEP regimens containing long-acting injectable cabotegravir or daily oral tenofovir disoproxil fumarate/emtricitabine among men who have sex with men in the United States: a mathematical modelling study for HPTN 083. Lancet regional health - Americas. 2023;18:100416. doi:10.1016/j.lana.2022.100416.

[84] Mitchell KM, Dimitrov D, Silhol R, Geidelberg L, Moore M, Liu A. The potential effect of COVID-19-related disruptions on HIV incidence and HIV-related mortality among men who have sex with men in the USA: a modelling study. Lancet HIV. 2021;8(4):e206–e215. doi:10.1016/S2352-3018(21)00022-9.

[85] Mitchell KM, Hoots B, Dimitrov D, German D, Flynn C, Farley JE, et al. Improvements in the HIV care continuum needed to meaningfully reduce HIV incidence among men who have sex with men in Baltimore, US: a modelling study for HPTN 078. Journal of the international AIDS society. 2019;22(3):e25246. doi:10.1002/jia2.25246.

[86] Mitchell KM, Prudden HJ, Washington R, Isac S, Rajaram SP, Foss AM, et al. Potential impact of pre-exposure prophylaxis for female sex workers and men who have sex with men in Bangalore, India: a mathematical modelling study. Journal of the international AIDS society. 2016;19(1):20942. doi:10.7448/IAS.19.1.20942.

[87] Mukandavire C, Walker J, Schwartz S, Boily MC, Danon L, Lyons C, et al. Estimating the contribution of key populations towards the spread of HIV in Dakar, Senegal. Journal of the international AIDS society. 2018;21(S5):e25126. doi:10.1002/jia2.25126.

[88] Reitsema M, Wallinga J, van Sighem AI, Bezemer D, van der Valk M, van Aar F, et al. Impact of Varying Pre-exposure Prophylaxis Programs on HIV and Neisseria gonorrhoeae Transmission Among MSM in the Netherlands: A Modeling Study. Journal of acquired immune deficiency syndromes. 2024;97(4):325–333. doi:10.1097/qai.0000000000003511.

[89] Robineau O, Velter A, Barin F, Boelle PY. HIV transmission and pre-exposure prophylaxis in a high risk MSM population: a simulation study of location-based selection of sexual partners. PLoS One. 2017;12(11):e0189002. doi:10.1371/journal.pone.0189002.

[90] Rozhnova G, Heijne JCM, Basten M, den Daas C, Matser A, Kretzschmar M. Impact of sexual trajectories of men who have sex with men on the reduction in HIV transmission by pre-exposure prophylaxis. Epidemics. 2019;28:100337. doi:10.1016/j.epidem.2019.03.003.

[91] Rozhnova G, van der Loeff MFS, Heijne JCM, Kretzschmar ME. Impact of heterogeneity in sexual behavior on effectiveness in reducing HIV transmission with test-and-treat strategy. PLOS computational biology. 2016;12(8):1–20. doi:10.1371/journal.pcbi.1005012.

[92] Silhol R, Maheu-Giroux M, Soni N, Fotso AS, Rouveau N, Vautier A, et al. The impact of past HIV interventions and diagnosis gaps on new HIV acquisitions, transmissions, and HIV-related deaths in Cote d’Ivoire, Mali, and Senegal. AIDS. 2024;38(12):1783–1793. doi:10.1097/QAD.0000000000003974.

[93] Silhol R, Baral S, Bowring AL, Mukandavire C, Njindam IM, Rao A, et al. Quantifying the evolving contribution of HIV interventions and key populations to the HIV epidemic in Yaoundé, Cameroon. Journal of acquired immune deficiency syndromes. 2021;86(4):396–405. doi:10.1097/QAI.0000000000002580.

[94] Silhol R, Geidelberg L, Mitchell KM, Mishra S, Dimitrov D, Bowring A, et al. Assessing the potential impact of disruptions due to COVID-19 on HIV among key and lower-risk populations in the largest cities of Cameroon and Benin. Journal of acquired immune deficiency syndromes. 2021;87(3):899–911. doi:10.1097/QAI.0000000000002663.

[95] Silhol R, Boily MC, Dimitrov D, German D, Flynn C, Farley JE, et al. Understanding the HIV epidemic among MSM in Baltimore: a modeling study estimating the impact of past HIV interventions and who acquired and contributed to infections. Journal of acquired immune deficiency syndromes. 2020;84(3):253–262. doi:10.1097/QAI.0000000000002340.

[96] Singleton AL, Marshall BDL, Zang X, Nunn AS, Goedel WC. Added benefits of pre-exposure prophylaxis use on HIV incidence with minimal changes in efficiency in the context of high treatment engagement among men who have sex with men. AIDS patient care STDS. 2020;34(12):506–515. doi:10.1089/apc.2020.0151.

[97] Shen M, Xiao Y, Rong L, Meyers LA, Bellan SE. Early antiretroviral therapy and potent second-line drugs could decrease HIV incidence of drug resistance. Proceedings of the royal society. 2017;284(1857). doi:10.1098/rspb.2017.0525.

[98] Stansfield SE, Herbeck JT, Gottlieb GS, Abernethy NF, Murphy JT, Mittler JE, et al. Test-and-treat coverage and HIV virulence evolution among men who have sex with men. Virus evolution. 2021;7(1):veab011. doi:10.1093/ve/veab011.

[99] Stone J, Mukandavire C, Boily MC, Fraser H, Mishra S, Schwartz S, et al. Estimating the contribution of key populations towards HIV transmission in South Africa. Journal of the international AIDS society. 2021;24(1):e25650. doi:10.1002/jia2.25650.

[100] Tollett Q, Safdar S, Gumel AB. Dynamics of a two-group model for assessing the impacts of pre-exposure prophylaxis, testing and risk behaviour change on the spread and control of HIV/AIDS in an MSM population. Infectious disease modelling. 2024;9(1):103–127. doi:10.1016/j.idm.2023.11.004.

[101] Vermeer W, Gurkan C, Hjorth A, Benbow N, Mustanski BM, Kern D, et al. Agent-based model projections for reducing HIV infection among MSM: Prevention and care pathways to end the HIV epidemic in Chicago, Illinois. PLoS One. 2022;17(10):e0274288. doi:10.1371/journal.pone.0274288.

[102] Viguerie A, Gopalappa C, Lyles CM, Farnham PG. The effects of HIV self-testing on HIV incidence and awareness of status among men who have sex with men in the United States: Insights from a novel compartmental model. Epidemics. 2024;49:100796. doi:10.1016/j.epidem.2024.100796.

[103] Wang Y, Tanuma J, Li J, Iwahashi K, Peng L, Chen C, et al. Elimination of HIV transmission in Japanese MSM with combination interventions. The Lancet regional health – Western Pacific. 2022;23. doi:10.1016/j.lanwpc.2022.100467.

[104] Wang L, Moqueet N, Simkin A, Knight J, Ma H, Lachowsky NJ, et al. Mathematical modelling of the influence of serosorting on the population-level HIV transmission impact of pre-exposure prophylaxis. AIDS. 2021;35(7):1113–1125. doi:10.1097/QAD.0000000000002826.

[105] Zang X, Piske M, Humphrey L, Enns B, Sui Y, Marshall BDL, et al. Estimating the epidemiological impact of reaching the objectives of the Florida integrated HIV prevention and care plan in Miami-Dade County. Lancet regional health - Americas. 2023;27:100623. doi:10.1016/j.lana.2023.100623.

[106] Zhang C, Webb GF, Lou J, Shepherd BE, Qian HZ, Liu Y, et al. Predicting the long-term impact of voluntary medical male circumcision on HIV incidence among men who have sex with men in Beijing, China. AIDS care. 2020;32(3):343–353. doi:10.1080/09540121.2019.1679704.

[107] Sweileh WM. Global research activity on mathematical modeling of transmission and control of 23 selected infectious disease outbreak. Globalization and health. 2022;18(1):4. doi:10.1186/s12992-022-00803-x.

[108] Anderle RV, de Oliveira RB, Rubio FA, Macinko J, Dourado I, Rasella D. Modelling HIV/AIDS epidemiological complexity: a scoping review of agent-based models and their application. PLoS One. 2024;19(2):e0297247. doi:10.1371/journal.pone.0297247.

[109] Eaton JW, Johnson LF, Salomon JA, Bärnighausen T, Bendavid E, Bershteyn A, et al. HIV treatment as prevention: systematic comparison of mathematical models of the potential impact of antiretroviral therapy on HIV incidence in South Africa. PLOS medicine. 2012;9(7):1–20. doi:10.1371/journal.pmed.1001245.

[110] Kloek M, Bulstra CA, van Noord L, Al-Hassany L, Cowan FM, Hontelez JAC. HIV prevalence among men who have sex with men, transgender women and cisgender male sex workers in sub-Saharan Africa: a systematic review and meta-analysis. Journal of the international AIDS society. 2022;25(11):e26022. doi:10.1002/jia2.26022.

[111] Stevens O, Sabin K, Anderson R, Garcia SA, Willis K, Rao A, et al. Population size, HIV prevalence, and antiretroviral therapy coverage among key populations in sub-Saharan Africa: collation and synthesis of survey data 2010-2023. medRxiv. 2024;doi:10.1101/2022.07.27.22278071.

[112] Okano JT, Robbins D, Palk L, Gerstoft J, Obel N, Blower S. Testing the hypothesis that treatment can eliminate HIV: a nationwide, population-based study of the Danish HIV epidemic in men who have sex with men. The Lancet infectious diseases. 2016;16(7):789–796. doi:10.1016/S1473-3099(16)30022-6.

[113] Bavinton B, Grulich A. HIV pre-exposure prophylaxis: scaling up for impact now and in the future. The Lancet public health. 2021;6. doi:10.1016/S2468-2667(21)00112-2.

[114] Moyo PL, Nunu WN. Exploring barriers and facilitators that influence uptake of oral pre-exposure prophylaxis among men who have sex with men in Bulawayo, Zimbabwe: key stakeholder’s perspectives. American journal of men’s health. 2024;18(1):15579883231223377. doi:10.1177/15579883231223377.

[115] Kurth AE, Celum C, Baeten JM, Vermund SH, Wasserheit JN. Combination HIV prevention: significance, challenges, and opportunities. Current HIV/AIDS Reports. 2011;8(1):62–72. doi:10.1007/s11904-010-0063-3.

[116] Iwuji CC, McGrath N, de Oliveira T, Porter K, Pillay D, Fisher M, et al. The art of HIV elimination: past and present science. Journal of AIDS and clinical research. 2015;6. doi:10.4172/2155-6113.1000525.

[117] Galvani AP, Pandey A, Fitzpatrick MC, Medlock J, Gray GE. Defining control of HIV epidemics. The Lancet HIV. 2018;5(11):e667–e670. doi:10.1016/S2352-3018(18)30178-4.

[118] Ghys PD, Williams BG, Over M, Hallett TB, Godfrey-Faussett P. Epidemiological metrics and benchmarks for a transition in the HIV epidemic. PLoS Med. 2018;15(10):e1002678. doi:10.1371/journal.pmed.1002678.

[119] Kapadia F, Landers S. Ending the HIV epidemic: getting to zero and staying at zero. American journal of public health. 2020;110(1):15–16. doi:10.2105/AJPH.2019.305462.

[120] Reitsema M, van Hoek AJ, van der Loeff MS, Hoornenborg E, van Sighem A, Wallinga J, et al. Preexposure prophylaxis for men who have sex with men in the Netherlands: impact on HIV and Neisseria gonorrhoeae transmission and cost-effectiveness. AIDS. 2020;34(4):621–630. doi:10.1097/QAD.0000000000002469.

[121] Jansen IA, Geskus RB, Davidovich U, Jurriaans S, Coutinho RA, Prins M, et al. Ongoing HIV-1 transmission among men who have sex with men in Amsterdam: a 25-year prospective cohort study. AIDS. 2011;25(4). doi:10.1097/qad.0b013e328342fbe9.

[122] Brooks RA, Landovitz RJ, Kaplan RL, Lieber E, Lee SJ, Barkley TW. Sexual risk behaviors and acceptability of HIV pre-exposure prophylaxis among HIV-negative gay and bisexual men in serodiscordant relationships: a mixed methods study. AIDS patient care STDS. 2011;26(2):87–94. doi:10.1089/apc.2011.0283.

[123] Schaefer R, Gregson S, Benedikt C. Widespread changes in sexual behaviour in eastern and southern Africa: Challenges to achieving global HIV targets? Longitudinal analyses of nationally representative surveys. Journal of the international AIDS society. 2019;22(6):e25329. doi:10.1002/jia2.25329.

[124] The Joint United Nations Programme on HIV/AIDS (UNAIDS). UNAIDS data 2023; 2023. Available from: https://www.unaids.org/sites/default/files/media_asset/data-book-2023_en.pdf.

[125] Santoso D, Asfia SKBM, Mello MB, Baggaley RC, Johnson CC, Chow EPF, et al. HIV prevalence ratio of international migrants compared to their native-born counterparts: A systematic review and meta-analysis. eClinicalMedicine. 2022;53. doi:10.1016/j.eclinm.2022.101661.

[126] Fiorentino M, Coulibaly B, Couderc C, Keita BD, Anoma C, Dah E, et al. Men who have sex with both men and women in West Africa: factors associated with a high behavioral risk of acquiring HIV from male partners and transmission to women (CohMSM ANRS 12324—Expertise France). Archives of sexual behavior. 2024;53(2):757–769. doi:10.1007/s10508-023-02715-2.

[127] Fiorentino M, Yanwou N, Gravier-Dumonceau Mazelier R, Eubanks A, Roux P, Laurent C, et al. Sexual behaviours and risk with women in MSM in sub-Saharan Africa. AIDS. 2024;38(3). doi:10.1097/QAD.0000000000003787.

[128] The Joint United Nations Programme on HIV/AIDS (UNAIDS). Global AIDS strategy 2021–2026: end inequalities, end AIDS; 2021. Available from: https://www.unaids.org/sites/default/files/media_asset/global-AIDS-strategy-2021-2026_en.pdf.

[129] Ayieko J, Petersen ML, Kamya MR, Havlir DV. PEP for HIV prevention: are we missing opportunities to reduce new infections? Journal of the international AIDS society. 2022;25(5):e25942. doi:10.1002/jia2.25942.

[130] Ribeiro Banze Á, Muleia R, Nuvunga S, Boothe M, Semá Baltazar C. Trends in HIV prevalence and risk factors among men who have sex with men in Mozambique: implications for targeted interventions and public health strategies. BMC public health. 2024;24(1):1185. doi:10.1186/s12889-024-18661-0.

[131] Lewin SR, Attoye T, Bansbach C, Doehle B, Dubé K, Dybul M, et al. Multi-stakeholder consensus on a target product profile for an HIV cure. The Lancet HIV. 2021;8(1):e42–e50. doi:10.1016/S2352-3018(20)30234-4.

