## Supplementary figures and tables for "Prospects of HIV elimination among men who have sex with men: a systematic review of modeling studies"

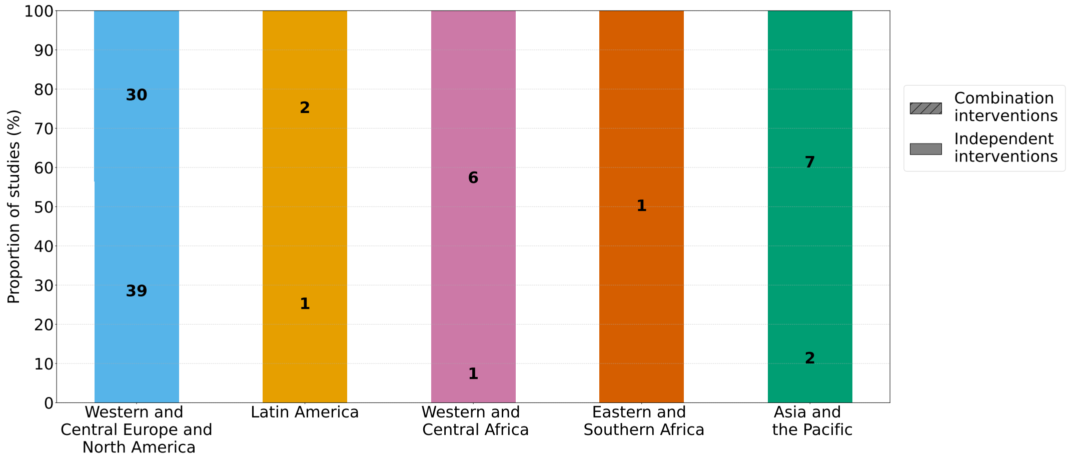


**Figure S1. Inclusion of modeled combination and individual interventions in studies.** We refer to individual interventions as interventions that were studied independently. Combination interventions imply that at least two interventions were studied in combination. The labels on each bar represent the number of times interventions were included in combination or independently.

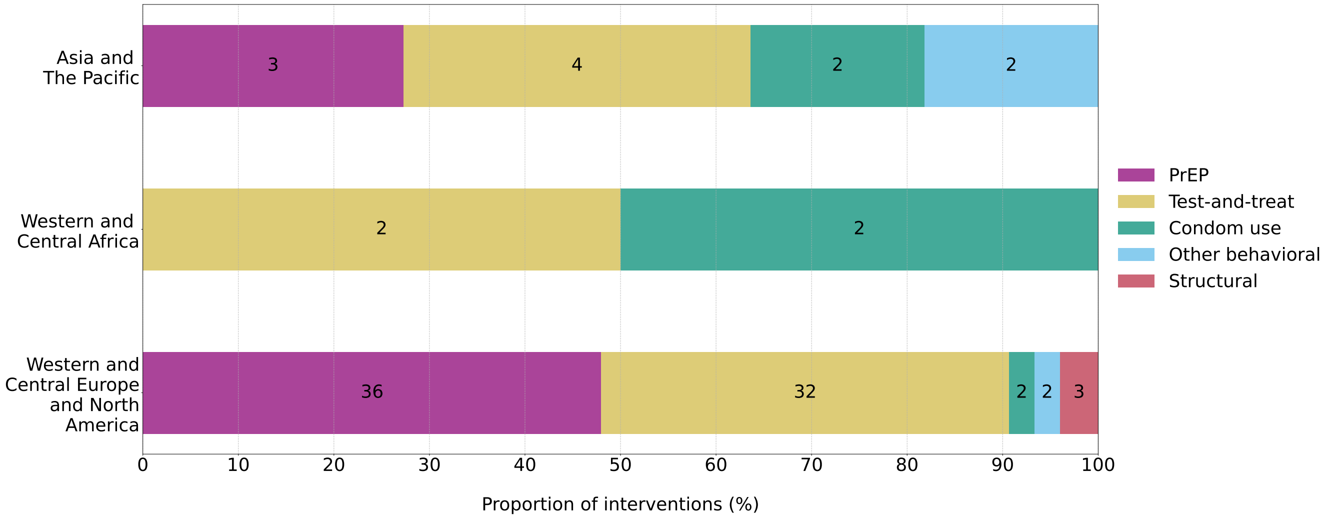


**Figure S2. Distribution of modeled interventions by UNAIDS region in studies that defined elimination.** The labels on each bar represent the number of times the intervention was included. A single study might include multiple interventions. Pre-exposure prophylaxis = PrEP.

881

**Table S1. Achievability of elimination through modeled combination and individual interventions.** ND = not discussed. - = not applicable. UNAIDS = The Joint United Nations Programme on HIV/AIDS.

| **Elimination achievable** | **Interventions** | **Feasibility** | **UNAIDS regions** | | | |
|---|---|---|---|---|---|---|
| | | | Western and Central Europe and North America | Western and Central Africa | Asia and The Pacific | **Total** |
| | | | n (%) | n (%) | n (%) | **n** (%) |
| Yes | Combination | Likely | 3 (13.64) | 0 (0.00) | 2 (66.67) | **5** (19.23) |
| | | Unlikely | 9 (40.91) | 0 (0.00) | 0 (0.00) | **9** (34.62) |
| | | ND | 10 (45.45) | 1 (100.00) | 1 (33.33) | **12** (46.15) |
| | | **Total** | **22** (73.33) | **1** (100.00) | **3** (75.00) | **26** (72.22) |
| | Individual | Likely | 0 (0.00) | 0 (0.00) | 1 (100.00) | **1** (10.00) |
| | | Unlikely | 8 (100.00) | 0 (0.00) | 0 (0.00) | **9*** (90.00) |
| | | ND | 0 (0.00) | 0 (0.00) | 0 (0.00) | **0** (0.00) |
| | | **Total** | **8** (26.67) | **0** (0.00) | **1** (25.00) | **10*** (27.78) |
| | **Total** | | **30** (71.43) | **1** (50.00) | **4** (80.00) | **36*** (72.00) |
| No | Combination | - | 9 (75.00) | 1 (100.00) | 0 (0.00) | **10** (71.43) |
| | Individual | - | 3 (25.00) | 0 (0.00) | 1 (100.00) | **4** (28.57) |
| | **Total** | | **12** (28.57) | **1** (50.00) | **1** (20.00) | **14** (28.00) |
| **Total** | | | **42** | **2** | **5** | **50***,† |

* The study [29] without the reference to a specific location predicted elimination as achievable, but unlikely, using an individual intervention.
† The study [82] was not included in this table because it defined elimination but did not directly assess its achievability.

**Table S2. Interventions included in modeled scenarios by achievability of elimination.** UNAIDS = The Joint United Nations Programme on HIV/AIDS.

| **Elimination achievable** | **Intervention** | **UNAIDS region** | | | |
|---|---|---|---|---|---|
| | | Western and Central Europe and North America | Western and Central Africa | Asia and The Pacific | **Total** |
| | | n (%) | n (%) | n (%) | **n** (%) |
| Yes | PrEP | 25 (44.64) | 0 (0.00) | 3 (30.00) | **28** (40.58) |
| | Test-and-treat | 24 (42.86) | 1 (50.00) | 3 (30.00) | **29*** (42.03) |
| | Condom use | 2 (3.57) | 1 (50.00) | 2 (20.00) | **5** (7.25) |
| | Other behavioral | 2 (3.57) | 0 (0.00) | 2 (20.00) | **4** (5.80) |
| | Structural | 3 (5.36) | 0 (0.00) | 0 (0.00) | **3** (4.35) |
| | **Total** | **56** (74.67) | **2** (50.00) | **10** (90.91) | **69*** (75.82) |
| No | PrEP | 11 (57.89) | 0 (0.00) | 0 (0.00) | **11** (50.00) |
| | Test-and-treat | 8 (42.11) | 1 (50.00) | 1 (100.00) | **10** (45.45) |
| | Condom use | 0 (0.00) | 1 (40.00) | 0 (0.00) | **1** (4.55) |
| | Other behavioral | 0 (0.00) | 0 (0.00) | 0 (0.00) | **0** (0.00) |
| | Structural | 0 (0.00) | 0 (0.00) | 0 (0.00) | **0** (0.00) |
| | **Total** | **19** (25.33) | **2** (50.00) | **1** (9.09) | **22** (24.18) |
| **Total** | | **75** | **4** | **11** | **91*** |

* The study [29] did not reference a specific location but did predict elimination as achievable using test-and-treat.
The study by Milwid et al. [82] defined elimination but did not assess any elimination scenarios and, therefore, was not counted in this table.

**Table S3. Achievability of elimination by type of modeled intervention.** UNAIDS = The Joint United Nations Programme on HIV/AIDS.

| **Intervention** | **Elimination achievable** | **UNAIDS region** | | | |
|---|---|---|---|---|---|
| | | Western and Central Europe and North America n (%) | Western and Central Africa n (%) | Asia and The Pacific n (%) | **Total** **n** (%) |
| PrEP | Yes | 25 (69.44) | 0 (0.00) | 3 (100.00) | **28** (71.79) |
| | No | 11 (30.56) | 0 (0.00) | 0 (0.00) | **11** (28.21) |
| | **Total** | **36** (48.00) | **0** (0.00) | **3** (27.27) | **39** (42.86) |
| Test-and-treat | Yes | 24 (75.00) | 1 (50.00) | 3 (75.00) | **29*** (74.36) |
| | No | 8 (25.00) | 1 (50.00) | 1 (25.00) | **10** (25.64) |
| | **Total** | **32** (42.67) | **2** (50.00) | **4** (36.36) | **39*** (42.86) |
| Condom use | Yes | 2 (100.00) | 1 (50.00) | 2 (100.00) | **5** (83.33) |
| | No | 0 (0.00) | 1 (50.00) | 0 (0.00) | **1** (16.67) |
| | **Total** | **2** (2.67) | **2** (50.00) | **2** (18.18) | **6** (6.59) |
| Other behavioral | Yes | 2 (100.00) | 0 (0.00) | 2 (100.00) | **4** (100.00) |
| | No | 0 (0.00) | 0 (0.00) | 0 (0.00) | **0** (0.00) |
| | **Total** | **2** (2.67) | **0** (0.00) | **2** (18.18) | **4** (4.40) |
| Structural | Yes | 3 (100.00) | 0 (0.00) | 0 (0.00) | **3** (100.00) |
| | No | 0 (0.00) | 0 (0.00) | 0 (0.00) | **0** (0.00) |
| | **Total** | **3** (4.00) | **0** (0.00) | **0** (0.00) | **3** (3.30) |
| **Total** | | **75** | **4** | **11** | **91*** |

* The study [29] did not reference a specific location but did predict elimination as achievable using test-and-treat.
The study by Milwid et al. [82] defined elimination but did not assess an elimination scenario and, therefore, was not counted in this table.

**Table S4. Comprehensiveness criteria.**

| Criterion | Considerations | Score considerations |
|---|---|---|
| Adherence to interventions | Does the model include parameters to account for adherence to the primary interventions? | 0: Not included for any primary interventions. 1: Included for some but not all primary interventions. 2: Included for all primary interventions. |
| Sexual risk compensation | Does the model include parameters to account for changes in sexual behavior as a result of interventions? | 0: Not included. 1: Included. |
| Openness of the MSM population | Does the model account for the introduction of new individuals to the population who have HIV? Does the model allow for mixing between MSM and other subpopulations, such as the heterosexual population? | 0: No introduction of individuals with HIV or mixing between MSM and other subpopulations. 1: At least one of these considerations included. |
| Uncertainty in model outcomes | Does the model report results with appropriate uncertainty, such as simulation intervals or uncertainty from parameter sets? Have any sensitivity analyses been performed with respect to parameters that were estimated or assumed? | 0: No sensitivity analyses and no uncertainty reported in the results. 1: One of these considerations included. 2: Both considerations included. |
| Model validation | Is there an attempt made to validate the model outputs against existing data? | 0: No attempt made. 1: Informal attempt made, i.e, details are unclear or validation performed against empirically derived targets. 2: Clear validation carried out. |

**Table S5. Model comprehensiveness scoring.**

| Authors (year) | Adherence to interventions | Sexual risk compensation | Openness of the MSM population | Uncertainty in model outcomes | Model validation | Overall score |
|---|---|---|---|---|---|---|
| Cambiano et al (2023) [26] | 2 | 0 | 1 | 2 | 0 | **5** |
| Chen et al (2022) [28] | 2 | 0 | 1 | 2 | 2 | **7** |
| David et al (2020) [29] | 0 | 0 | 0 | 1 | 0 | **1** |
| Fojo et al (2021) [35] | 2 | 0 | 1 | 2 | 0 | **5** |
| Gilmour et al (2020)* [38] | 1 | 0 | 0 | 2 | 0 | **3** |
| Goedel et al (2020) [39] | 2 | 0 | 0 | 2 | 0 | **4** |
| Goedel et al (2018) [40] | 2 | 0 | 0 | 2 | 0 | **4** |
| Le Guillou et al (2021) [42] | 2 | 1 | 0 | 1 | 0 | **4** |
| Gutowska et al (2022) [44] | 2 | 0 | 0 | 2 | 0 | **4** |
| Hamilton et al (2023) [18] | 2 | 0 | 1 | 1 | 0 | **4** |
| Hamilton et al (2018) [48] | 2 | 0 | 1 | 2 | 0 | **5** |
| Hansson et al (2020) [49] | 2 | 0 | 0 | 2 | 0 | **4** |
| Hansson et al (2019)* [50] | 2 | 0 | 0 | 2 | 0 | **4** |
| Jenness et al (2020) [55] | 2 | 1 | 0 | 2 | 1 | **6** |
| Jenness et al (2019) [56] | 2 | 1 | 0 | 2 | 1 | **6** |
| Jijon et al (2021) [59] | 2 | 1 | 0 | 2 | 0 | **5** |
| Khanna et al (2021) [64] | 2 | 0 | 1 | 2 | 0 | **5** |
| Khanna et al (2019) [65] | 2 | 0 | 1 | 2 | 0 | **5** |
| Lee et al (2023) [69] | 2 | 0 | 1 | 2 | 1 | **6** |
| Lee et al (2022) [70] | 2 | 0 | 1 | 2 | 1 | **6** |
| Lima et al (2021) [71] | 2 | 1 | 0 | 2 | 2 | **7** |
| Lou et al (2017) [74] | 2 | 1 | 0 | 2 | 0 | **5** |
| Lu et al (2020) [76] | 2 | 0 | 0 | 2 | 2 | **6** |
| Maheou-Giroux et al (2017) [77] | 2 | 0 | 1 | 2 | 0 | **5** |
| Martin et al (2020) [81] | 2 | 0 | 0 | 1 | 1 | **4** |
| Massey et al (2023) [80] | 1 | 0 | 1 | 0 | 0 | **2** |
| Milwid et al (2022) [82] | 2 | 0 | 0 | 1 | 2 | **5** |
| Mitchell et al (2023) [83] | 2 | 0 | 0 | 2 | 2 | **6** |
| Mitchell et al (2019) [85] | 2 | 0 | 0 | 2 | 2 | **6** |
| Mitchell et al (2016)* [86] | 2 | 1 | 1 | 1 | 2 | **7** |
| Palk et al (2018)* [4] | 2 | 0 | 1 | 2 | 0 | **5** |
| Rozhnova et al (2018)* [16] | 2 | 0 | 0 | 2 | 0 | **4** |
| Rozhnova et al (2016) [91] | 2 | 0 | 0 | 2 | 0 | **4** |
| Silhol et al (2021) [93] | 2 | 0 | 1 | 2 | 1 | **6** |
| Singleton et al (2020) [96] | 2 | 0 | 0 | 2 | 0 | **4** |
| Tollett et al (2024) [100] | 1 | 0 | 0 | 2 | 2 | **5** |
| Vermeer et al (2022) [101] | 2 | 0 | 0 | 2 | 2 | **6** |
| Wang et al (2022)* [103] | 1 | 0 | 0 | 2 | 0 | **3** |
| Zang et al (2023) [105] | 1 | 0 | 1 | 0 | 2 | **4** |

* These studies predicted elimination in the modeled intervention scenarios, which were also discussed as being plausible in a real-world context.

**Table S6. Comprehensiveness score distribution by model type.**

| Score | **Model type** | | | |
|---|---|---|---|---|
| | ABM | DCM | SCM | **Total** |
| | n (%) | n (%) | n (%) | n (%) |
| 1 | 0 (0.00) | 1 (5.00) | 0 (0.00) | **1** (2.56) |
| 2 | 0 (0.00) | 0 (0.00) | 1 (25.00) | **1** (2.56) |
| 3 | 0 (0.00) | 2 (10.00) | 0 (0.00) | **2** (5.13) |
| 4 | 5 (33.33) | 6 (30.00) | 1 (25.00) | **12** (30.77) |
| 5 | 5 (33.33) | 4 (20.00) | 2 (50.00) | **11** (28.21) |
| 6 | 5 (33.33) | 4 (20.00) | 0 (0.00) | **9** (23.08) |
| 7 | 0 (0.00) | 3 (15.00) | 0 (0.00) | **3** (7.69) |
| **Total** | **15** | **20** | **4** | **39** |
| **Average score** | **5.00** | **4.80** | **4.00** | **4.79** |
